# Bern Sleep-Wake Registry: temporal extraction of comorbidities from unstructured clinical text using a large language model

**DOI:** 10.64898/2026.10.25.26364496

**Authors:** Irina Filchenko, Fabio Dennstädt, Kseniia Zub, Felix I. Pompizii, Julia van der Meer, Antoine Adamantidis, Nikola Cihoric, Markus H. Schmidt, Claudio L.A. Bassetti

**Affiliations:** Department of Neurology, Bern University Hospital (Inselspital) and University of Bern, 3010, Bern, Switzerland; Department of Radiation Oncology, Inselspital, Bern University Hospital and University of Bern, Bern, 3010, Switzerland; Department of Digital Medicine, Inselspital, Bern University Hospital and University of Bern, 3010, Bern, Switzerland; Data Semantics and Decision Making Institute (DSDMI), 9000, St. Gallen, Switzerland; Medical Faculty, University of Bern, 3010, Bern, Switzerland

**Author notes:** <u>Corresponding author:</u> Irina Filchenko, MD, PhD, Department of Neurology, Bern University Hospital (Inselspital) and University of Bern, 3010, Bern, Switzerland. These authors contributed equally to this work (Irina Filchenko, Fabio Dennstädt).

**Keywords:** large language models, natural language processing, temporal information, electronic health records, clinical phenotyping, clinical informatics, real-world data

## Abstract

**Introduction:** Large language models (LLMs) may enable the extraction of clinically relevant information from unstructured electronic health records, but most previous work focused on identifying diagnoses without preserving their temporal context. We evaluated the ability of an LLM to extract diagnoses and associated dates from longitudinal clinical reports.

**Methods:** In an observational study using the Bern Sleep-Wake Registry, diagnostic annotations from 248 patients comprising 2343 clinical entries were analyzed. Six common comorbidity groups, subdivided into 32 hierarchical diagnostic categories, were evaluated. Using gpt-oss-20b, we extracted diagnosis presence, the number of diagnostic instances, and dates per instance. A reference standard was established through independent review by two physicians, with disagreements adjudicated by a third physician. LLM performance was assessed using diagnostic classification and temporal agreement. Direct annotation costs were compared.

**Results:** At the patient level, the LLM achieved an overall F1 score of 0.88 for diagnosis presence and 0.96 for the presence of a reported date. For the earliest reported diagnosis date, agreement with the reference standard was 86.73% at the day level. When considering the cumulative set of dates for each diagnosis, date recall was 87.63%, the extra-date rate was 20.88%, and full agreement was achieved in 58.19% of cases. Human annotators showed substantial disagreement in temporal annotation, with a date recall of 75.49%, an extra-date rate of 17.89%, and full agreement in 58.64% of cases. LLM-based extraction reduced direct annotation costs by 94.65%.

**Conclusions:** LLMs can extract diagnoses and their dates from heterogeneous clinical text with high agreement with an adjudicated reference standard. These findings support the scalable use of LLMs for longitudinal clinical registries.

## Introduction

Real-world data in healthcare are defined as data relating to patient health status and/or the delivery of healthcare that are routinely collected from a variety of sources [1]. Although randomized clinical trials remain the highest level of evidence for evaluating treatment efficacy, real-world data provide insights into routine clinical practice and improve the generalizability [2].

To characterize clinical management and outcomes, particularly in case of heterogeneous or unclear disease trajectories, patient registries provide systematic, longitudinal data (e.g., European Narcolepsy database or Swiss Clinical Quality Management registry for rheumatic diseases) [3,4]. Patient registries are often based on information from electronic health records (EHRs). While export of structured fields is possible in some EHRs (e.g., classification codes with the date of entry), a substantial amount of clinically relevant information is documented in unstructured free-text clinical reports (e.g., annotations to the classification codes). Consequently, many important details are not accessible using conventional *a priori*-defined databases, whereas manual extraction of features on demand is not feasible in large-scale registries.

Large language models (LLM) represent a promising solution to address this issue. LLMs were used to successfully extract structured information from unstructured clinical text across different medical domains as well as data types, including categorical (e.g., medication extraction with an F1=0.99% [5], boolean questions on oncologic trials with an F1=0.92 [6]) and continuous variables (e.g., medication dosage with F1-score of 84.98% [5], and clinical scores with accuracy of 87.69% [7]).

One important application of LLMs for information extraction is the identification of comorbidities from unstructured clinical text. For example, we previously demonstrated that the instruction-tuned Mistral-24B model achieved a macro classification accuracy of 95% and an F1 score of 92% across six common comorbidity categories [8]. Similarly, Zhang et al. (2025) reported 100% accuracy and a 98.5% F1 score for extracting symptoms [5]. However, most existing studies focused on static information extraction without preserving their temporal context.

In clinical practice and research, the temporal context of diagnoses is essential. The evolution of comorbidities provides critical information for risk stratification and prognostic modelling. Importantly, the date on which a diagnosis is entered into the EHR does not necessarily reflect the actual time of disease onset or diagnosis. Although some EHR systems provide a dedicated field for the date of diagnosis, this field is often missing, especially for chronic conditions such as diabetes or hypertension. Consequently, registry studies frequently rely on the date of diagnosis entry [9,10]. However, this timestamp reflects when the diagnosis was documented rather than when the condition was first identified. In an analysis comparing a cancer registry date and a claims-based index date for cancer diagnosis, the index date was identical to the cancer registry date for 47% of patients and preceded the registry date for 28% of patients [10] .

Despite the importance of temporal information, extracting temporal structures from unstructured clinical text remains challenging and is insufficiently explored in natural language processing. We addressed this gap by investigating the ability of an LLM to extract diagnoses and their dates from unstructured clinical reports using the Bern Sleep-Wake Registry (BSWR), a large real-world registry from a tertiary sleep center.

## Methods

### Study population

We conducted a registry-based observational study using data from the BSWR, a longitudinal clinical registry of patients evaluated at the Sleep-Wake-Epilepsy Center of the Inselspital, University Hospital Bern, Switzerland. The BSWR is implemented as a REDCap database with relevant information (e.g., demographics) linked to the raw data of the objective sleep–wake assessments stored on the local server (Van der Meer et al., submitted).

Patients were eligible for scientific evaluation if they provided general consent. The BWSR was approved by the local ethics committee (Kantonale Ethikkommission Bern [KEK]-Nr. 2022-00415), ensuring compliance with the Human Research Act (HRA) and Ordinance on Human Research with the Exception of Clinical Trials (HRO).

### Dataset

Diagnostic information was obtained from the ‘I-pdos’ EHR (CompuGroup Medical Schweiz AG, Switzerland) covering the period from 2009-05-28 to 2024-03-18. For the present comparison between LLM and ground truth (**Figure 1A**), diagnostic annotations consisting of a structured diagnosis-entry dates and the corresponding free-text entries were extracted for a randomly selected sample of 248 patients [8]. Each diagnostic record for an individual patient was split into pairs based on its text and date forming multiple individual entries. The structured entry date was deliberately excluded from the extraction task to ensure that the model was evaluated on its ability to recover temporal information from the clinical text itself. The resulting dataset comprised 2343 entries from 248 patients, with a mean±SD of 9.44±7.51 entries per patient (maximum: 34).

**Figure 1.**
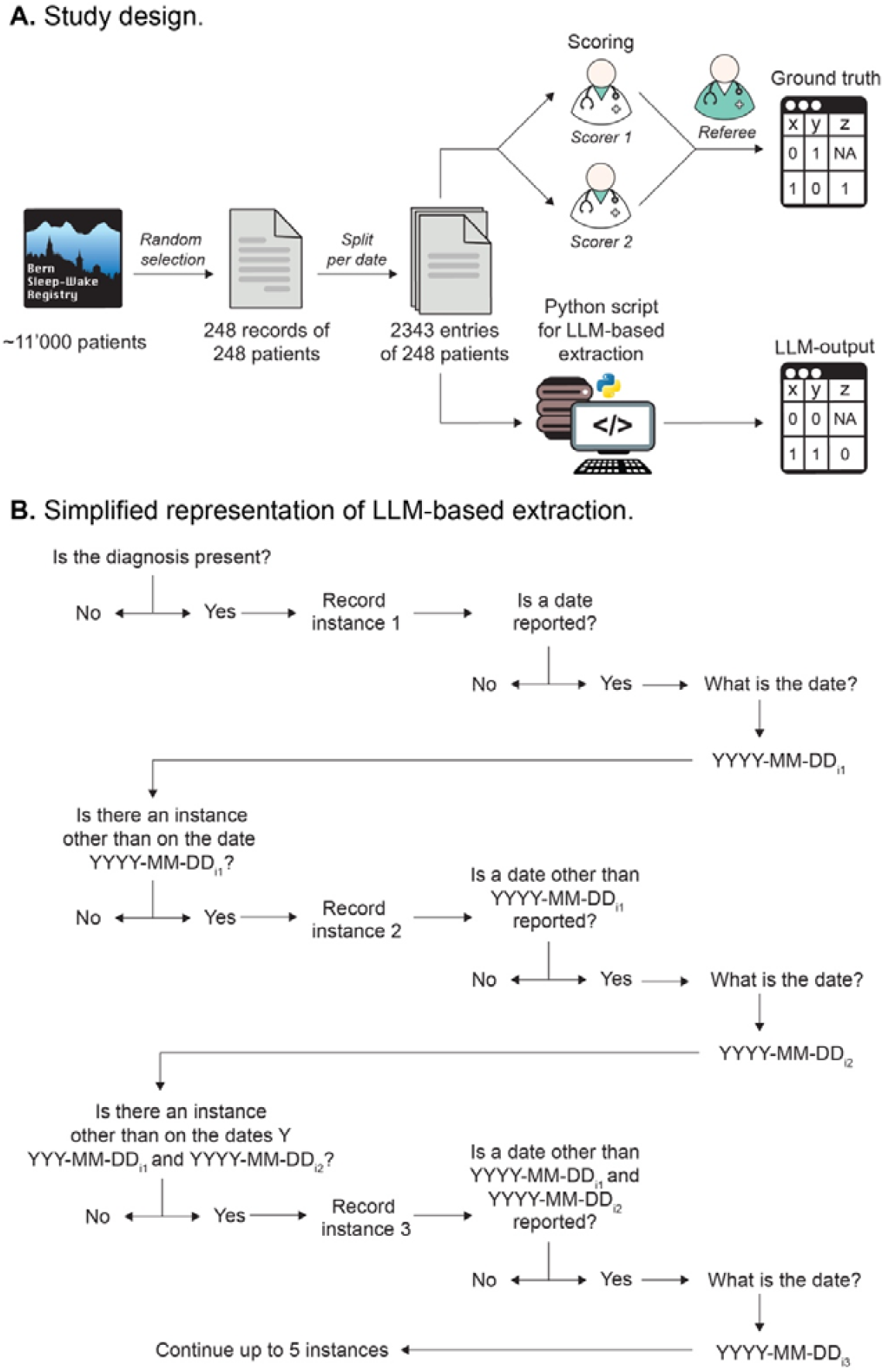
**A**. Study design. A random sample of 248 patients was selected, yielding 2343 individual diagnostic entries. Two independent physicians performed manual annotation, with disagreements adjudicated by a third reviewer to establish the ground truth. A Python script orchestrated the LLM-based extraction across the same set of entries. **B**. Simplified representation of the LLM extraction for a single diagnosis. For each diagnostic entry, the model first determined whether a target diagnosis was present. If present, the model recorded the first diagnostic instance and assessed whether a date could be extracted from the clinical text. The model then iteratively checked for additional diagnostic instances occurring on distinct dates.

For the analysis, we selected six common comorbidities based on the International Classification of Diseases, 11th Revision (ICD-11). These categories were subdivided into 32 hierarchical diagnostic subcategories spanning up to four levels of the diagnostic hierarchy (**Supplementary Table 1**).

**Table 1.** Selected parameters of LLM performance versus ground truth at the record level (n=248).

| Index | Diagnosis | Diagnosis present |  |  |  |  |  |  |  |  | Date reported |  |  |  |  |  |  |  |  |
| --- | --- | --- | --- | --- | --- | --- | --- | --- | --- | --- | --- | --- | --- | --- | --- | --- | --- | --- | --- |
|  |  | TP | TN | FP | FN | Acc. | Recall | Spec. | F1 | Kappa | TP | TN | FP | FN | Acc. | Recall | Spec. | F1 | Kappa |
| 1 | Ischemic stroke or transient ischemic attack | 37 | 207 | 4 | 0 | 0.98 | 1.00 | 0.98 | 0.95 | 0.98 | 36 | 1 | 0 | 0 | 1.00 | 1.00 | 1.00 | 1.00 | 1.00 |
| 1.1 | Transient ischaemic attack | 12 | 233 | 3 | 0 | 0.99 | 1.00 | 0.99 | 0.89 | 0.99 | 10 | 2 | 0 | 0 | 1.00 | 1.00 | 1.00 | 1.00 | 1.00 |
| 1.1.1 | Amaurosis fugax | 1 | 247 | 0 | 0 | 1.00 | 1.00 | 1.00 | 1.00 | 1.00 | 1 | 0 | 0 | 0 | 1.00 | 1.00 | - | 1.00 | - |
| 1.2 | Cerebral ischaemic stroke | 31 | 216 | 1 | 0 | 1.00 | 1.00 | 1.00 | 0.98 | 1.00 | 30 | 0 | 1 | 0 | 0.97 | 1.00 | 0.00 | 0.98 | 0.00 |
| 1.2.1 | Cerebral ischaemic stroke due to extracranial large artery atherosclerosis | 3 | 243 | 0 | 2 | 0.99 | 0.60 | 1.00 | 0.75 | 0.99 | 3 | 0 | 0 | 0 | 1.00 | 1.00 | - | 1.00 | - |
| 1.2.2 | Cerebral ischaemic stroke due to intracranial large artery atherosclerosis | 1 | 245 | 2 | 0 | 0.99 | 1.00 | 0.99 | 0.50 | 0.99 | 1 | 0 | 0 | 0 | 1.00 | 1.00 | - | 1.00 | - |
| 1.2.3 | Cerebral ischaemic stroke due to embolic occlusion | 15 | 229 | 2 | 2 | 0.98 | 0.88 | 0.99 | 0.88 | 0.98 | 15 | 0 | 0 | 0 | 1.00 | 1.00 | - | 1.00 | - |
| 1.2.4 | Cerebral ischaemic stroke due to small artery occlusion | 6 | 241 | 1 | 0 | 1.00 | 1.00 | 1.00 | 0.92 | 1.00 | 5 | 0 | 0 | 1 | 0.83 | 0.83 | - | 0.91 | 0.00 |
| 1.2.5 | Cerebral ischaemic stroke due to other known cause | 5 | 238 | 4 | 1 | 0.98 | 0.83 | 0.98 | 0.67 | 0.98 | 3 | 0 | 1 | 1 | 0.60 | 0.75 | 0.00 | 0.75 | -0.11 |
| 2 | Diabetes | 19 | 222 | 6 | 1 | 0.97 | 0.95 | 0.97 | 0.84 | 0.97 | 14 | 4 | 1 | 0 | 0.95 | 1.00 | 0.80 | 0.97 | 0.87 |
| 2.1 | Type 1 diabetes mellitus | 1 | 247 | 0 | 0 | 1.00 | 1.00 | 1.00 | 1.00 | 1.00 | 1 | 0 | 0 | 0 | 1.00 | 1.00 | - | 1.00 | - |
| 2.2 | Type 2 diabetes mellitus | 17 | 227 | 3 | 1 | 0.98 | 0.94 | 0.99 | 0.89 | 0.98 | 12 | 3 | 1 | 1 | 0.88 | 0.92 | 0.75 | 0.92 | 0.72 |
| 3 | Dyslipidemia | 48 | 194 | 6 | 0 | 0.98 | 1.00 | 0.97 | 0.94 | 0.97 | 23 | 18 | 3 | 4 | 0.85 | 0.85 | 0.86 | 0.87 | 0.79 |
| 3.1 | Hyperlipoproteinaemia | 10 | 226 | 9 | 3 | 0.95 | 0.77 | 0.96 | 0.63 | 0.95 | 3 | 4 | 0 | 3 | 0.70 | 0.50 | 1.00 | 0.67 | 0.63 |
| 3.1.1 | Hypercholesterolaemia | 10 | 228 | 7 | 3 | 0.96 | 0.77 | 0.97 | 0.67 | 0.96 | 3 | 4 | 0 | 3 | 0.70 | 0.50 | 1.00 | 0.67 | 0.63 |
| 3.1.2 | Hypertriglyceridaemia | 0 | 244 | 4 | 0 | 0.98 | - | 0.98 | - | 0.98 | - | - | - | - | - | - | - | - | - |
| 3.2 | Hypolipoproteinaemia | 0 | 248 | 0 | 0 | 1.00 | - | 1.00 | - | 1.00 | - | - | - | - | - | - | - | - | - |
| 4 | Hypertension | 67 | 177 | 3 | 1 | 0.98 | 0.99 | 0.98 | 0.97 | 0.98 | 37 | 23 | 5 | 2 | 0.90 | 0.95 | 0.82 | 0.91 | 0.84 |
| 4.1 | Essential hypertension | 2 | 214 | 32 | 0 | 0.87 | 1.00 | 0.87 | 0.11 | 0.87 | 1 | 1 | 0 | 0 | 1.00 | 1.00 | 1.00 | 1.00 | 1.00 |
| 4.2 | Hypertensive heart disease | 14 | 227 | 5 | 2 | 0.97 | 0.88 | 0.98 | 0.80 | 0.97 | 9 | 4 | 0 | 1 | 0.93 | 0.90 | 1.00 | 0.95 | 0.87 |
| 4.3 | Hypertensive renal disease | 2 | 246 | 0 | 0 | 1.00 | 1.00 | 1.00 | 1.00 | 1.00 | 2 | 0 | 0 | 0 | 1.00 | 1.00 | - | 1.00 | - |
| 4.4 | Pulmonary heart disease or diseases of pulmonary circulation | 6 | 236 | 3 | 3 | 0.98 | 0.67 | 0.99 | 0.67 | 0.98 | 5 | 0 | 0 | 1 | 0.83 | 0.83 | - | 0.91 | 0.00 |
| 4.1.1 | Pulmonary thromboembolism | 2 | 243 | 2 | 1 | 0.99 | 0.67 | 0.99 | 0.57 | 0.99 | 2 | 0 | 0 | 0 | 1.00 | 1.00 | - | 1.00 | - |
| 4.1.2 | Pulmonary hypertension | 4 | 242 | 0 | 2 | 0.99 | 0.67 | 1.00 | 0.80 | 0.99 | 3 | 0 | 0 | 1 | 0.75 | 0.75 | - | 0.86 | 0.00 |
| 5 | Atrial fibrillation | 13 | 233 | 2 | 0 | 0.99 | 1.00 | 0.99 | 0.93 | 0.99 | 13 | 0 | 0 | 0 | 1.00 | 1.00 | - | 1.00 | - |
| 5.1 | Paroxysmal atrial fibrillation | 9 | 239 | 0 | 0 | 1.00 | 1.00 | 1.00 | 1.00 | 1.00 | 9 | 0 | 0 | 0 | 1.00 | 1.00 | - | 1.00 | - |
| 5.2 | Persistent atrial fibrillation | 1 | 245 | 2 | 0 | 0.99 | 1.00 | 0.99 | 0.50 | 0.99 | 1 | 0 | 0 | 0 | 1.00 | 1.00 | - | 1.00 | - |
| 5.3 | Permanent atrial fibrillation | 0 | 247 | 1 | 0 | 1.00 | - | 1.00 | - | 1.00 | - | - | - | - | - | - | - | - | - |
| 6 | Sleep-disordered breathing | 136 | 109 | 3 | 0 | 0.99 | 1.00 | 0.97 | 0.99 | 0.98 | 100 | 32 | 3 | 1 | 0.97 | 0.99 | 0.91 | 0.98 | 0.93 |
| 6.1 | Central sleep apnoeas | 14 | 229 | 5 | 0 | 0.98 | 1.00 | 0.98 | 0.85 | 0.98 | 11 | 2 | 0 | 1 | 0.93 | 0.92 | 1.00 | 0.96 | 0.78 |
| 6.2 | Obstructive sleep apnoea | 105 | 119 | 24 | 0 | 0.90 | 1.00 | 0.83 | 0.90 | 0.88 | 81 | 20 | 4 | 0 | 0.96 | 1.00 | 0.83 | 0.98 | 0.90 |
| 6.3 | Sleep-related hypoventilation or hypoxemia disorders | 3 | 235 | 8 | 2 | 0.96 | 0.60 | 0.97 | 0.38 | 0.96 | 3 | 0 | 0 | 0 | 1.00 | 1.00 | - | 1.00 | - |
|  | Overall | 594 | 7176 | 142 | 24 | 0.98 | 0.96 | 0.98 | 0.88 | 0.98 | 437 | 118 | 19 | 20 | 0.93 | 0.96 | 0.86 | 0.96 | 0.84 |

FN – false-negative, FP – false-positive, TN – true-negative, TP – true-positive.

For each diagnostic category, the extraction task allowed up to five distinct diagnostic instances per patient (**Figure 1B**). A diagnostic instance was defined as the occurrence of the same diagnosis at a distinct date. For example, a stroke documented on 8 July 2016 and a subsequent stroke documented on 1 February 2026 were considered two separate instances. If the date of a diagnostic instance was not available, the diagnosis was considered present but its date was classified as unknown. Thus, a diagnosis with an unknown date and the same diagnosis documented with a known date were treated as separate instances where applicable.

Furthermore, the dates were available with different level of precision, and the uniform format was required. If the day, month, and year were available, the date was recorded in the format YYYY-MM-DD. If only the month and year were known, the date was recorded as YYYY-MM-01. If only the year was known, the date was recorded as YYYY-01-01.

### Large language model–based information extraction

Information was extracted from the diagnostic annotations using gpt-oss-20b [11] and a Python-based library for LLM-based information extraction (data-element-extractor, v. 0.3.0; **Figure 2**) [6]. This LLM was selected since it was among the most capable open-source LLMs at the time the study (i.e., July 2026; the rationale is detailed in **Supplementary material 1**). Given the rapid pace of technological development and the fact that a thorough comparison of different models was conducted previously [12–15], we did not aim to perform an additional model comparison.

**Figure 2.**
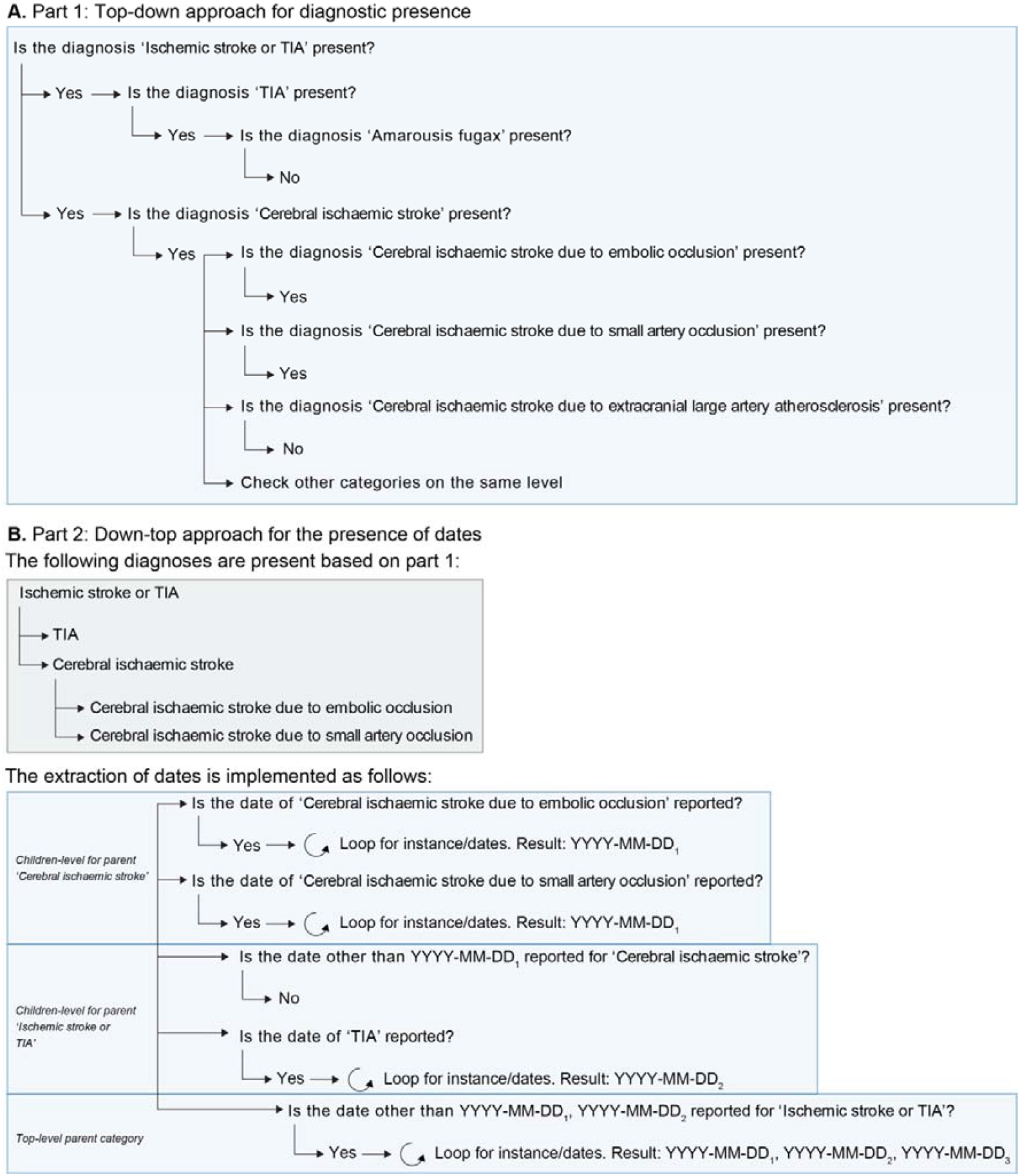
Two-stage approach for diagnostic presence and date extraction. **A**. Top-down approach for determining the presence of diagnoses within the hierarchical diagnostic structure. Parent categories are assessed first, followed by progressively more specific child diagnoses if the parent category is present. **B**. Down-top approach for extracting dates from diagnoses identified as presented in Part 1. Dates are first extracted at the child level, with duplicate dates being excluded. Additional dates reported only at higher levels of the hierarchy are subsequently extracted. This approach allows dates to be assigned to the most specific applicable diagnostic level while retaining dates reported at broader diagnostic levels.

The complete extraction prompt is provided in **Supplementary material 2**. In brief, for each entry, the model was instructed to identify the presence of diagnoses based on diagnostic categories and subcategories. The model was not provided with the ground-truth annotations during inference. For each identified diagnosis, the model extracted (1) whether the diagnosis was present, (2) the number of distinct diagnostic instances, and (3) the date associated with each diagnostic instance when a date could be determined from the diagnostic annotation.

### Ground truth

A reference standard was established by independent manual review of the diagnostic annotations by two physicians. Consistent with the LLM output, for each predefined diagnostic category, the annotators assessed whether the diagnosis was present and, if present, recorded the number of diagnostic instances and the date of each instance when this could be determined from the clinical information. Disagreements between the annotators were reviewed by a third physician, who acted as a referee and established the ground truth. The annotation was done via a web-based interface developed by the Data Semantics and Decision Making Institute (Switzerland).

### Statistical analysis

Statistical analyses were performed using MATLAB 2024b. LLM output was compared with ground truth at two levels: the patient level, based on information consolidated across all 2343 entries from 248 patients, and the entry level, based on individual entries. The patient-level analysis reflected the clinical task of identifying the emergence of a diagnosis across a longitudinal record and reduced the influence of repeated documentation of the same information. The entry-level analysis provided a granular assessment of model performance.

At the patient level, a diagnosis or diagnosis date was considered present if reported in at least one entry. Diagnostic presence and date reporting were evaluated as binary outcomes, with date reporting assessed only when the diagnosis was correctly identified. Agreement with ground truth was assessed using accuracy, sensitivity (recall), specificity, precision, F1 score, and Cohen’s κ. For cases in which a diagnosis date was correctly identified as present, date agreement was assessed at the day, month, and year levels. The primary date outcome was exact agreement of the earliest reported diagnosis date at the patient level. Additional analyses assessed the agreement of all unique reported dates using exact-date recall, extra-date rate, and complete exact-date agreement (**Supplementary Material 3**).

At the entry level, diagnostic presence was assessed using the same approach. For date analyses, only leaf-level diagnostic categories were included to avoid double-counting diagnoses represented by hierarchical parent and child categories. We assessed the number of reported dates per entry and agreement in the number of dates, followed by set-based agreement of unique dates using the same metrics as in the patient-level analysis.

## Results

### Agreement on the presence of diagnoses (boolean categories)

The analysis of agreement by disease category, including comparisons between the LLM and the ground truth and between human scorers, is provided in **Supplementary Material 4**.

#### Human scorers

In the analysis of 248 consolidated patient records, inter-scorer agreement was high (F1=0.93, κ=0.99). A low F1 was observed for ischemic stroke due to other known etiology (0.44). No true positive classifications were observed for ischemic stroke due to extracranial large artery atherosclerosis, ischemic stroke due to intracranial large artery atherosclerosis, pulmonary thromboembolism, and sleep-related hypoventilation or hypoxemia disorders. Agreement with the ground truth was excellent for both Scorer 1 (F1=0.96, κ=0.99) and Scorer 2 (F1=0.97, κ=1.00).

In the analysis of the 2343 diagnostic entries, interscorer agreement among human scorers was excellent, with an overall κ=1.00. Substantial variability was observed across diagnostic categories. In particular, the F1 was low for essential hypertension (0.40) and ischemic stroke due to other known etiology (0.10), largely owing to false-negative classifications. No true positive classifications were observed for pulmonary thromboembolis, ischemic stroke due to extracranial large artery atherosclerosis, or ischemic stroke due to intracranial large artery atherosclerosis. These findings suggest that certain diagnostic categories present considerable challenges for human scorers.

#### LLM vs. ground truth

In the analysis of 248 consolidated patient records **(Table 1)**, the LLM demonstrated high overall agreement with the ground truth for diagnosis presence (F1=0.88). Performance was consistent across diagnostic categories, with five of 32 having near-perfect agreement (F1≥0.95). Lower performance was observed for essential hypertension (F1=0.11) and sleep-related hypoventilation or hypoxemia disorders (F1=0.38), driven primarily by false-positive classifications. These results were consistent to the analysis of underlying 2343 diagnostic entries (F1=0.88; **Table 2**), showing high variability across diagnoses.

**Table 2.** Selected parameters of LLM performance versus ground truth at the entry level (n=2343). Further parameters are detailed in **Supplementary material 2**.

| Index | Diagnosis | Presence of a diagnosis (yes/no) |  |  |  |  |  |  |  |  | Number of dates reported<sup>1</sup> |  |
| --- | --- | --- | --- | --- | --- | --- | --- | --- | --- | --- | --- | --- |
|  |  | TP | TN | FP | FN | Acc. | Recall | Spec. | F1 | Kappa | N | Agreement<sup>2</sup>, % |
| 1 | Ischemic stroke or transient ischemic attack | 116 | 2217 | 6 | 4 | 1.00 | 0.97 | 1.00 | 0.96 | 1.00 | 0 | - |
| 1.1 | Transient ischaemic attack | 21 | 2318 | 4 | 0 | 1.00 | 1.00 | 1.00 | 0.91 | 1.00 | 18 | 88.89 |
| 1.1.1 | Amaurosis fugax | 3 | 2340 | 0 | 0 | 1.00 | 1.00 | 1.00 | 1.00 | 1.00 | 3 | 66.67 |
| 1.2 | Cerebral ischaemic stroke | 99 | 2236 | 2 | 6 | 1.00 | 0.94 | 1.00 | 0.96 | 1.00 | 38 | 86.84 |
| 1.2.1 | Cerebral ischaemic stroke due to extracranial large artery atherosclerosis | 6 | 2331 | 0 | 6 | 1.00 | 0.50 | 1.00 | 0.67 | 1.00 | 6 | 100.00 |
| 1.2.2 | Cerebral ischaemic stroke due to intracranial large artery atherosclerosis | 1 | 2338 | 4 | 0 | 1.00 | 1.00 | 1.00 | 0.33 | 1.00 | 1 | 100.00 |
| 1.2.3 | Cerebral ischaemic stroke due to embolic occlusion | 33 | 2304 | 2 | 4 | 1.00 | 0.89 | 1.00 | 0.92 | 1.00 | 33 | 81.82 |
| 1.2.4 | Cerebral ischaemic stroke due to small artery occlusion | 16 | 2323 | 3 | 1 | 1.00 | 0.94 | 1.00 | 0.89 | 1.00 | 16 | 87.50 |
| 1.2.5 | Cerebral ischaemic stroke due to other known cause | 9 | 2321 | 8 | 5 | 0.99 | 0.64 | 1.00 | 0.58 | 0.99 | 9 | 66.67 |
| 2 | Diabetes | 57 | 2277 | 8 | 1 | 1.00 | 0.98 | 1.00 | 0.93 | 1.00 | 6 | 66.67 |
| 2.1 | Type 1 diabetes mellitus | 8 | 2335 | 0 | 0 | 1.00 | 1.00 | 1.00 | 1.00 | 1.00 | 8 | 100.00 |
| 2.2 | Type 2 diabetes mellitus | 39 | 2295 | 8 | 1 | 1.00 | 0.98 | 1.00 | 0.90 | 1.00 | 39 | 89.74 |
| 3 | Dyslipidemia | 117 | 2219 | 7 | 0 | 1.00 | 1.00 | 1.00 | 0.97 | 1.00 | 81 | 82.72 |
| 3.1 | Hyperlipoproteinaemia | 11 | 2304 | 16 | 12 | 0.99 | 0.48 | 0.99 | 0.44 | 0.99 | 0 | - |
| 3.1.1 | Hypercholesterolaemia | 11 | 2306 | 14 | 12 | 0.99 | 0.48 | 0.99 | 0.46 | 0.99 | 11 | 90.91 |
| 3.1.2 | Hypertriglyceridaemia | 0 | 2339 | 4 | 0 | 1.00 | - | 1.00 | - | 1.00 | 0 | - |
| 3.2 | Hypolipoproteinaemia | 0 | 2343 | 0 | 0 | 1.00 | - | 1.00 | - | 1.00 | 0 | - |
| 4 | Hypertension | 155 | 2175 | 6 | 7 | 0.99 | 0.96 | 1.00 | 0.96 | 0.99 | 68 | 86.76 |
| 4.1 | Essential hypertension | 9 | 2281 | 53 | 0 | 0.98 | 1.00 | 0.98 | 0.25 | 0.98 | 9 | 55.56 |
| 4.2 | Hypertensive heart disease | 28 | 2307 | 5 | 3 | 1.00 | 0.90 | 1.00 | 0.88 | 1.00 | 28 | 89.29 |
| 4.3 | Hypertensive renal disease | 4 | 2339 | 0 | 0 | 1.00 | 1.00 | 1.00 | 1.00 | 1.00 | 4 | 100.00 |
| 4.4 | Pulmonary heart disease or diseases of pulmonary circulation | 6 | 2325 | 8 | 4 | 0.99 | 0.60 | 1.00 | 0.50 | 0.99 | 0 | - |
| 4.1.1 | Pulmonary thromboembolism | 2 | 2334 | 6 | 1 | 1.00 | 0.67 | 1.00 | 0.36 | 1.00 | 2 | 100.00 |
| 4.1.2 | Pulmonary hypertension | 4 | 2335 | 1 | 3 | 1.00 | 0.57 | 1.00 | 0.67 | 1.00 | 4 | 100.00 |
| 5 | Atrial fibrillation | 33 | 2307 | 3 | 0 | 1.00 | 1.00 | 1.00 | 0.96 | 1.00 | 5 | 100.00 |
| 5.1 | Paroxysmal atrial fibrillation | 24 | 2319 | 0 | 0 | 1.00 | 1.00 | 1.00 | 1.00 | 1.00 | 24 | 75.00 |
| 5.2 | Persistent atrial fibrillation | 2 | 2339 | 2 | 0 | 1.00 | 1.00 | 1.00 | 0.67 | 1.00 | 2 | 100.00 |
| 5.3 | Permanent atrial fibrillation | 0 | 2341 | 2 | 0 | 1.00 | - | 1.00 | - | 1.00 | 0 | - |
| 6 | Sleep-disordered breathing | 340 | 1984 | 3 | 16 | 0.99 | 0.96 | 1.00 | 0.97 | 0.99 | 22 | 95.45 |
| 6.1 | Central sleep apnoeas | 24 | 2310 | 6 | 3 | 1.00 | 0.89 | 1.00 | 0.84 | 1.00 | 24 | 75.00 |
| 6.2 | Obstructive sleep apnoea | 213 | 2033 | 83 | 14 | 0.96 | 0.94 | 0.96 | 0.81 | 0.96 | 213 | 87.32 |
| 6.3 | Sleep-related hypoventilation or hypoxemia disorders | 8 | 2318 | 10 | 7 | 0.99 | 0.53 | 1.00 | 0.48 | 0.99 | 8 | 62.50 |
| Overall |  | 1399 | 73193 | 274 | 110 | 0.99 | 0.93 | 1.00 | 0.88 | 0.99 | 682 | 85.48 |

<sup>1</sup>Only leaf cases were analyzed to avoid the bias from aggregation. <sup>2</sup>To YYYY-MM-DD. FN – false-negative, FP – false-positive, TN – true-negative, TP – true-positive.

We examined the LLM-reasoning for the diagnostic categories with F1<0.50 for the individual entries. The LLM tended to classify cases as essential hypertension when no secondary cause was explicitly mentioned, even when the available information did not demonstrate that secondary causes were adequately excluded (**Supplementary Material 5**).

For ischemic stroke due to intracranial atherosclerosis (four false positives, F1=0.33), one case appears to result from the LLM interpreting posterior stroke attributed to macroangiopathy as intracranial rather than extracranial large-artery stenosis (**Supplementary Material 6**, example 1). The other three cases involved atherosclerosis of the V4 arterial segment, which should be classified as intracranial [16], supporting the correctness of the LLM.

For pulmonary thromboembolism (six false positives, F1=0.33), the misclassification was likely due to an oversight from the scorers (**Supplementary Material 7**).

For the 16 false-positive classifications of hyperlipoproteinaemia (F1=0.44), all cases were initially classified as negative by both scorers and therefore were not included in the ground-truth assessment. Upon additional review, all 16 cases were positive for dyslipidemia (**Supplementary Material 8**).

For sleep-related hypoventilation or hypoxemia (F1=0.48), among 10 false-positives, five cases were found to be negative upon additional review, suggesting a scorers’ error, whereas in another five cases the discrepancy seemingly reflected differences in reporting (**Supplementary Material 9**). Seven entries classified as false-negative cases belonged to two patients (**Supplementary Material 10**). In the cases involving BiPAP therapy, the false-negative classification was well-supported (i.e., BiPAP increases lung volumes and improves oxygenation [17]), whereas the reasoning of LLM was deemed correct for the remaining cases.

### Agreement on the presence or absence of dates

The analysis of agreement by disease category, including comparisons between the LLM and the ground truth and between human scorers, is provided in **Supplementary Material 4**.

#### Human scorers

In the analysis of 248 consolidated patient records, boolean agreement on the presence of dates was high between the human scorers (F1=0.94, κ=0.79). The lowest F1 was observed for essential hypertension (0.67).

At the level of the 2343 diagnostic entries, the leaf-node analysis of true-positive cases assessed the number of dates per entry. The scorers agreed on the number of dates in 88.20% of cases. The lowest agreement was observed for transient ischaemic attack (66.67%; n=16).

#### LLM vs. ground truth

In the analysis of 248 consolidated patient records, the overall boolean agreement on the presence of dates between the LLM and the ground truth was high (F1=0.86, **Table 1**). The lowest F1 was observed for hypercholesterolaemia (0.67), driven primarily by false-positive classifications.

At the level of the 2343 diagnostic entries, the leaf-node analysis of true-positive cases assessed the number of dates per entry (**Table 2**). Overall, 85.48% entries showed a full agreement on the number of dates between the LLM and the ground truth. The lowest agreement was observed for essential hypertension (55.56%; n=9) and sleep-related hypoventilation or hypoxemia disorders (62.50%; n=8).

Review of the LLM reasoning indicated that, for essential hypertension and sleep-related hypoventilation or hypoxemia disorders, the LLM occasionally interpreted each mention of the diagnosis (e.g., during follow-up examinations or treatment controls) as a separate date leading to the overestimation (**Supplementary Materials 11 and 12**).

### Agreement on the reported dates

The analysis of agreement by disease category, including comparisons between the LLM and the ground truth and between human scorers, is provided in **Supplementary Material 4**.

#### Human scorers

In the analysis of 248 consolidated patient records, the interscorer agreement for the earliest date was 75.25% on the day level, 79.17% on the month level, and 90.44% on the year level (**Figure 3**). No agreement for the earliest date was observed in two cases of cerebral ischaemic stroke due to other known cause. The lowest day-level agreement for the earliest date was observed for essential hypertension (4.29%, n=286). Day-level agreement of 100.00% for the earliest date was observed for four disease categories, whereas eight disease categories had 100.00% agreement at the year level. Complete match of the set of multiple dates per record was observed for 58.64% of records on the day level (**Figure 4A**).

**Figure 3.**
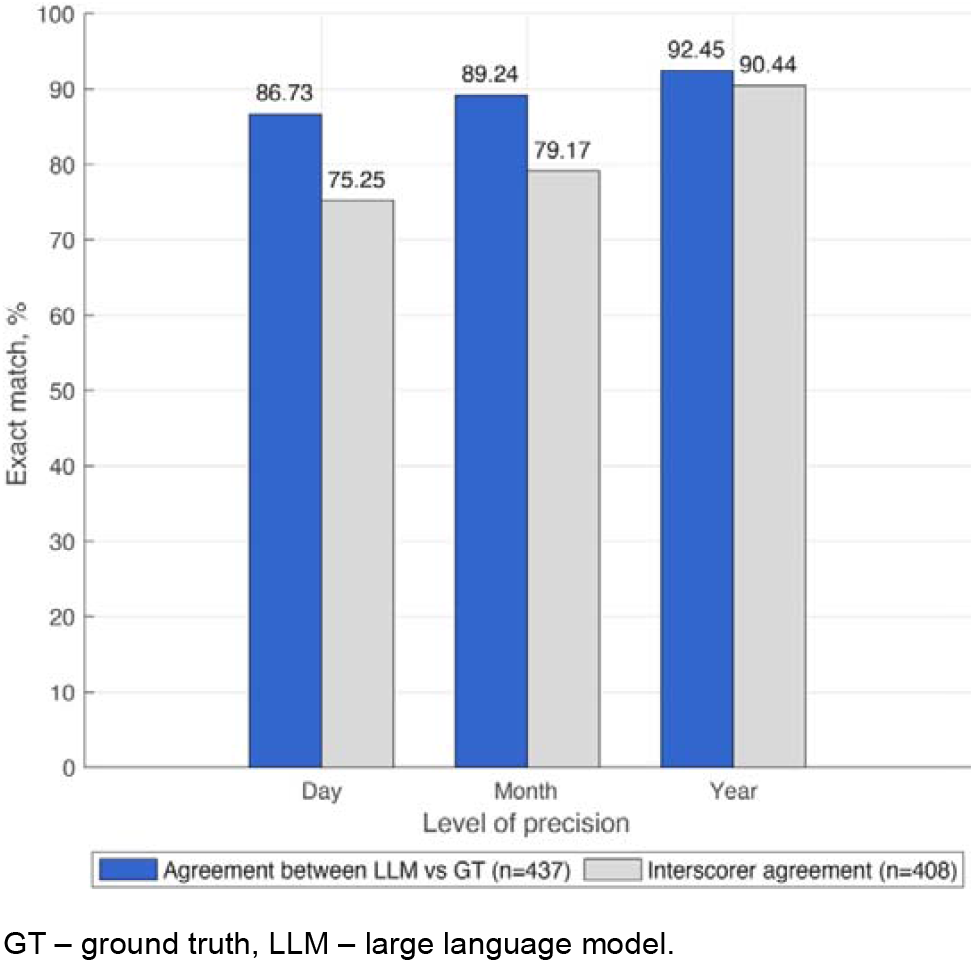
Record-level agreement for earliest date of diagnosis (n=248): cumulative statistics across 32 diseases. The description per disease category is detailed in **Supplementary material 2**.

**Figure 4.**
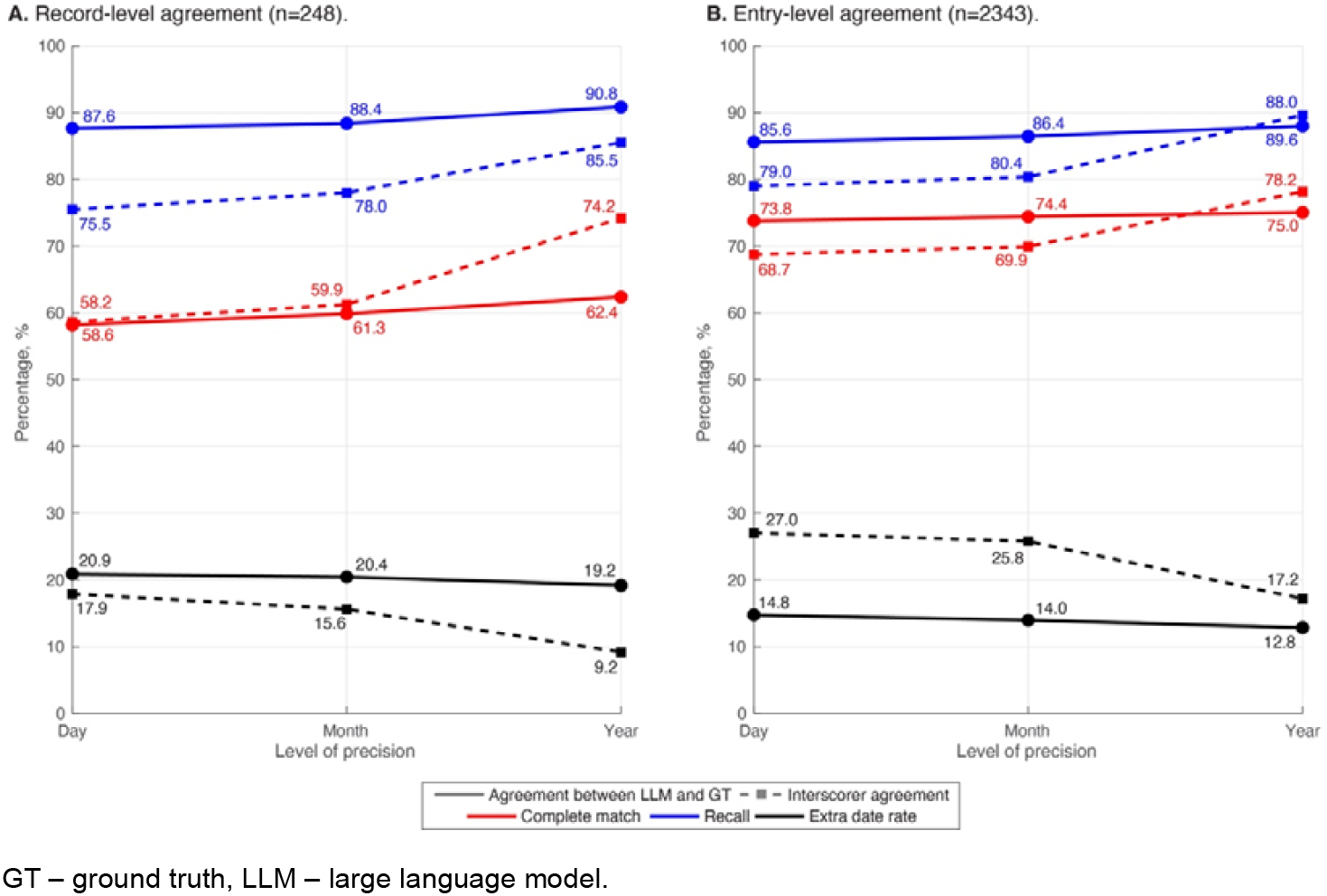
Record-level (**A**) and entry-level (**B**) agreement for the date of diagnosis: cumulative statistics across 32 diseases. The agreement is presented at the day-, month- and year-level of precision. The description per disease category is detailed in **Supplementary material 2**.

Among the 2343 diagnostic entries, the leaf-node analysis showed a day-level recall of 79.00% with 27.04% extra date rates (**Figure 4B;** Scorer 1: 92.04% and 8.98%, respectively; Scorer 2: 81.40% and 13.91%, respectively). Complete match was observed for 68.74% of records, with the lowest values in diabetes (50.00%, n=4), hypertension (51.06%, n=47), and dyslipidemia (52.73%, n=55). Two disease categories achieved 100% agreement.

#### LLM vs. ground truth

In the analysis of 248 consolidated patient records, the agreement for the earliest date was 86.73 on the day level, 89.24% on the month level, and 92.45% on the year level (**Figure 3**). The lowest day-level agreement for the earliest date was observed for hypertensive renal disease (50.00%, n=2) and cerebral ischaemic stroke due to small artery occlusion (60.00%, n=5). At the year level, >80% agreement for the earliest date was achieved across all disease categories except arterial hypertension (50%). Day-level agreement of 100.00% for the earliest date was observed for 16 disease categories, whereas 19 disease categories achieved 100.00% agreement at the year level. Complete match of the set of multiple dates per record was observed for 58.19% of records on the day level (**Figure 4A**).

Among the 2343 underlying diagnostic entries, the leaf-node analysis showed a day-level recall of 85.59% with 14.75% extra date rate (**Figure 4B**). Complete match was observed for 73.79% of records, with the lowest values in diabetes (33.33%, n=3) and essential hypertension (50.00%, n=8). Six disease categories achieved 100.00% agreement. The stereotypical reasoning for dates is detailed in **Supplementary material 13**.

### Estimation of direct annotation costs

The extraction was performed on a laboratory computer, for which electricity costs are difficult to estimate reliably unless measured directly at the source. Therefore, we estimated the direct computational cost of the annotation based on a cloud GPU rate of approximately CHF 2.00 per hour, representative of rates available in the region. At this rate, running the LLM for 96.0 hours on one GPU resulted in an estimated cost of CHF 192.00, compared with CHF 3588.00 for human annotation (69 person-hours at CHF 52.00 per hour: 12 and 30 hours per two scorers and 27 hours for the referee). This corresponds to an estimated cost saving of CHF 3396.00 (94.65%). The cost per unit of performance was also substantially lower for the LLM, at CHF 218.18 versus CHF 3718.13 per diagnosis F1 point and CHF 2.21 versus CHF 41.31per percentage point of date matching.

## Discussion

We evaluated the ability of an LLM to extract diagnostic information from real-world clinical data, with emphasis on the identification of diagnoses and their dates. A key novelty is the detailed assessment of LLM performance in identifying and reporting the dates of diagnoses across multiple diagnostic categories.

In line with previous studies demonstrating the use of LLMs for phenotyping (F1 ranging from 0.72 0.95) [8,18–20], we found high agreement between the LLM and the ground truth for diagnostic presence, with an F1=0.88 at both the patient and entry level. Importantly, the agreement between the LLM and ground truth was slightly lower than the interscorer agreement (i.e., patient-level: F1=0.93; entry-level: F1=0.94). Detailed review of the discordant cases revealed the limitations in the ground truth rather than incorrect LLM reasoning. For example, the medication for hyperlipoproteinaemia was overseen by the scorers in all 16 cases classified false-positively by the LLM. This observation highlights an ability of LLM to identify diagnoses implicitly supported by contextual information. At the same time, LLM may overinterpret the evidence. This issue was apparent for essential hypertension (F1=0.25), which is a diagnosis of exclusion implying an evaluation of secondary causes (e.g., sleep-related breathing disorder) [21]. In clinical practice, such an evaluation is rarely performed extensively [22] and essential hypertension may be documented as the presumed diagnosis, which is also reflected in the low entry-level interscorer agreement (F1=0.40). The LLM occasionally reproduced this pattern by classifying cases as essential hypertension based on the absence of an explicitly documented secondary cause.

A central contribution of this study was evaluating the ability of an LLM to extract diagnosis dates from real-world free clinical text. The LLM performed well in identifying whether a date was reported both at the patient level (F1=0.96 for LLM vs. ground truth, interscorer F1=0.94) and at the entry level (exact agreement for the number of dates: 85.48% for LLM vs. ground truth, 88.20% between scorers). Furthermore, the LLM correctly identified the earliest reported date in 86.73% of cases at the patient level, compared with 75.25% interscorer agreement. Performance improved as temporal precision decreased, suggesting that many discrepancies concerned the exact day rather than the broader temporal placement of the diagnosis. When considering the complete set of dates (i.e., not only the earliest date), the proportion of cases on the patient level in which all dates matched the ground truth at the day level was substantially lower and similar for the LLM and human scorers (≈58%). These findings indicate that LLMs can recover temporal information from clinical documentation, although the interpretation of this performance requires consideration of the inherent ambiguity of temporal annotation, as observed in human annotations.

Our findings are consistent with previous work demonstrating the feasibility of LLM-based temporal information extraction from clinical texts. Abdullahi et al. reported variable LLM performance versus physician-validated timestamps, underscoring the difficulty of assigning temporal information [23]. The TimeX pipeline achieved a timestamp-extraction accuracy of 81.24% in 102 manually annotated clinical notes, outperforming five baseline approaches by >14.86% [24]. Although broadly comparable, TimeX extracted the onset of individual phenotypes from clinical notes, whereas our study evaluated the reconstruction of multiple diagnostic instances across longitudinal data. Similarly, the PMOA-TTS corpus demonstrated the feasibility of large-scale timeline extraction using LLMs, with more than 5.6 million timestamps extracted from case reports [25]. However, case reports generally provide more structured and narratively coherent descriptions of disease courses than real-world registry data. Our study therefore extends this evidence by evaluating temporal extraction in free clinical texts characterized by repeated documentation, multiple comorbidities, and a variable temporal precision.

Extracting temporal information from free clinical texts provides an opportunity to reconstruct clinically meaningful trajectories without an extensive manual review. Given the high diagnostic and date recall observed in our study, an LLM could serve as a first-pass extraction tool for large-scale registry enrichment, with human review concentrated on challenging records. Such a semi-automated workflow could reduce the burden of data curation while preserving expert oversight.

The cost estimation further supports the scalability of LLM-based extraction, corresponding to a 94.65% reduction in direct annotation costs. Although these figures do not capture the full operational costs of developing a clinical NLP system, they illustrate the advantage of automated extraction with a comparable performance between the LLM and highly qualified human annotators. In practice, large-scale annotation is not always performed by physicians and may be delegated to less specialized personnel (e.g., master students) increasing the efficiency of LLM-based approaches. Chen et al. showed that a LLM outperforms attending physicians less often than unspecified medical doctors and medical students [26]. We envision the most appropriate application as semi-automated pipeline, in which the LLM performs large-scale extraction whereas human experts review uncertain cases. Importantly, information supporting the LLM predictions, including reasoning, should be available to reviewers to facilitate adjudication of ambiguous cases and provide transparency into the extraction.

The strengths of the study include a detailed, multilevel evaluation of LLM performance across key components of temporal extraction (i.e., diagnosis identification, detection of whether a date was reported, and reconstruction of the reported date itself). Since each textual entry has an entry date, the framework allows the entry date to be used as an alternative timestamp when no explicit diagnosis date is available. Furthermore, the reference was established through high-quality annotation by qualified medical reviewers with independent verification. We additionally assessed the reasoning of LLM, which, to our knowledge, was not evaluated in previous work and may be particularly valuable for supporting human review of ambiguous cases.

Several limitations should be acknowledged. First, the study included 32 diagnoses and data from a single tertiary sleep center, which may limit generalizability. Second, some diagnostic subcategories contained few cases, indicating the need of a careful interpretation of the performance. Third, defining a diagnostic instance remains challenging. The current framework extracts the dates of diagnoses but does not determine whether these dates may be interpreted as a disease onset, recurrence, or subsequent documentation of an established condition, as observed for essential hypertension. Future approaches should prioritize clinically meaningful events rather than every date associated with a diagnosis. This is particularly relevant for chronic or recurrent conditions such as multiple sclerosis, where repeated documentation of a persistent diagnosis does not necessarily indicate a new disease event. Finally, some discrepancies between the LLM and the reference standard appeared to reflect limitations of human annotations. Thus, disagreement with the reference does not necessarily indicate an incorrect LLM output and highlights the complexity of a temporal reference.

In conclusion, this study demonstrates that an LLM can extract diagnostic and temporal information from unstructured clinical reports with high agreement with an adjudicated reference. Our findings indicate that LLMs can be applied to large-scale real-world retrospective data that is difficult to structure at scale. Future work should focus on rich temporal representations (e.g., distinguishing disease episode from follow-up) and human-in-the-loop workflows that combine the scalability of LLM-based extraction with expert review of ambiguous cases.

## Data Availability

The analyzed datasets are not publicly available due to Swiss data protection laws. Data sharing may be considered upon request to the corresponding author, subject to the official agreements and compliance with Swiss data protection laws. The underlying code for this study is not publicly available but may be made available to qualified researchers on reasonable request from the corresponding author.

## Funding and acknowledgements

The data curation of the Bern Sleep-Wake Registry was supported by the Inselspital Grant “A digital reference network platform for clinical and experimental neuroscience – deep phenotyping and data integration” and the “Eurostars Project E!12034 SPAS”.

## Author contribution

IF and FD conceptualized the data extraction approach and developed the code for data extraction. IF performed the statistical analyses and drafted the manuscript, including Figures and Tables. KZ and FP independently performed the annotation, with IF serving as the adjudicator in cases of disagreement. JVDM curated the data for analysis. JVDM, MHS, and CLAB secured funding and administrated the project. All authors critically reviewed and approved the final manuscript.

## Consent to publish

All authors approved the final manuscript.

## Consent to participate

The Bern Sleep-Wake Registry was approved by the local ethics committee (Kantonale Ethikkommission Bern [KEK]-Nr. 2022-00415), ensuring compliance with the Human Research Act (HRA) and Ordinance on Human Research with the Exception of Clinical Trials (HRO).

## Data availability

The analyzed datasets are not publicly available due to Swiss data protection laws. Data sharing may be considered upon request to the corresponding author, subject to the official agreements and compliance with Swiss data protection laws.

## Code availability

The underlying code for this study is not publicly available but may be made available to qualified researchers on reasonable request from the corresponding author.

## Code availability

Not applicable

