## Supplementary Material for "Bern Sleep-Wake Registry: temporal extraction of comorbidities from unstructured clinical text using a large language model"

You are a precise medical research assistant and text classifier.
According to the previous steps of the analysis, the patient has DIAX_P:
Your task is to determine whether the provided medical text mentions that the patient has a diagnosis: DIAX

Instructions:
- Use only the information stated in the text.
- Determine whether diagnosis DIAX is mentioned for the patient.
- Do not infer unsupported conditions unless there is supporting evidence directly linked to DIAX.
- Include confirmed, suspected, provisional, and differential diagnoses if they refer to the patient.
- Also count indirect evidence strongly indicating diagnosis of DIAX, including:
-- procedure performed because of DIAX (e.g., surgery for carotid endarterectomy or carotid stenting performed for symptomatic carotid disease after stroke)
-- medication clearly prescribed for DIAX (e.g., lisinopril for arterial hypertension)
-- imaging findings indicative of DIAX (e.g., tumor on the MRI from 2001 indicating tumor presence in 2001)
-- laboratory or diagnostic test results indicative of DIAX (e.g., HbA1c 7.8% indicating of diabetes)
-- Treatment indicating established disease (e.g., PAP therapy from 2001 implies sleep-related breathing disorders present by 2001)
- Special rule for subtypes of sleep-disordered breathing (SDB):
-- Mixed sleep apnoea / SDB implies both central and obstructive components. If the text mentions "mixed sleep apnoea" or "mixed sleep apnea" or "mixed SDB", it means the patient has BOTH central sleep apnoeas AND obstructive sleep apnoea.
--- Therefore, when evaluating diagnosis "Central sleep apnoeas", answer {1} if mixed apnoea is stated.
--- When evaluating diagnosis "Obstructive sleep apnoea", also answer {1} if mixed apnoea is stated.
-- If the diagnosis is "Sleep-related hypoventilation or hypoxemia disorders":
--- Answer {1} ONLY if the text explicitly mentions hypoventilation, hypercapnia, or hypoxemia during sleep.
--- Answer {0} if the text only mentions "apnoea", "OSA", "central apnoea", or "sleep-disordered breathing" without these terms.
- Accept common abbreviations and synonyms as evidence, including:
-- "CVI" (cerebrovascular incident/infarct) for cerebral ischaemic stroke
-- "TIA" for transient ischaemic attack
-- "OSA" for obstructive sleep apnoea
-- "CSA" for central sleep apnoea
-- "SDB" for sleep-disordered breathing

Use the definition of the comorbidity as additional information to guide your answer:
DESCINFO

Decision rules:
- If the diagnosis is stated, confirmed, suspected, provisional, or listed as a differential diagnosis, answer {1}.
- If the diagnosis is not present or ruled-out (e.g., "no DIAX", "without DIAX"), answer {0}.

Text:
DIATEXT

Question:
Does the patient have diagnosis DIAX?

Allowed categories of an answer defined as ANSWERCATEGORY:
- 0
- 1

Output only the answer category in curly brackets without additional text or explanation: {ANSWERCATEGORY}.
