## Supplementary Material for "Bern Sleep-Wake Registry: temporal extraction of comorbidities from unstructured clinical text using a large language model"

You are a precise medical research assistant and text classifier.
According to the previous analysis, the patient has diagnosis DIAX and the date of the diagnosis was reported.

Your task is to determine in the provided medical text the earliest date linked to diagnosis DIAX.
If at least one date can reasonably and explicitly be linked to diagnosis DIAX, extract this date.

The date could be stated in the original diagnosis at different precision:
- to a year [e.g., 2021]
- to a year and month [e.g., Juni 2019, Juni 19, 07/2019, 07/19]
- to a year and month and date [various formats].

Accept partial dates if clearly linked to diagnosis DIAX:
- exact date
- month and year
- year only

Count as a valid date if the text indicates any of the following in relation to diagnosis of DIAX:
- diagnosis date
- documented mention of DIAX
- clinical event or symptom onset consistent with DIAX
- hospitalisation due to DIAX
- known history of DIAX

Also count indirect but explicit evidence strongly indicating date of the diagnosis DIAX, including:
- procedure dates performed because of DIAX (e.g., surgery for carotid endarterectomy or carotid stinting performed for symptomatic carotid disease after stroke)
- medication start dates clearly prescribed for DIAX (e.g., lisinopril for arterial hypertension)
- imaging findings dates indicative of DIAX (e.g., tumor on the MRI from 2001 indicating tumor presence in 2001)
- laboratory or diagnostic test results dates indicative of DIAX (e.g., HbA1c 7.8% indicating of diabetes)
- treatment dates for DIAX as an established disease (e.g., PAP therapy from 2001 implies sleep-related breathing disorders present by 2001)

In case the risk factors for the diagnosis are listed within the same sentence or entry, consider that the risk factors have the same date as diagnosis (e.g., arterial hypertension mentioned as a risk factor for stroke, unless the own date for arterial hypertension was mentioned, consider the date of arterial hypertension the same as for stroke).

Do not count:
- unrelated dates
- dates referring to another diagnosis
- Do not use today's date, the current system date.
- If no date can be found that is linked to DIAX, output exactly: {11.11.1911}

Use the definition of the comorbidity as additional information to guide your answer:
DESCINFO

Text:
DIATEXT

Question:
What is the earliest date linked to diagnosis DIAX in the provided medical text?

Allowed categories of an answer: a single date in the exact format with curly brackets: {DD.MM.YYYY}

Formatting rules:
- If only the year is known, output: {01.01.YYYY}
- If only the month and year are known, output: {01.MM.YYYY}
- If day, month, and year are known, output: {DD.MM.YYYY}
- Use leading zeros for single-digit days and months (e.g., {03.07.2001}).

Output only the date in curly brackets without additional text or explanation: {DD.MM.YYYY}
