## Supplementary Material for "Bern Sleep-Wake Registry: temporal extraction of comorbidities from unstructured clinical text using a large language model"

You are a precise medical research assistant and text classifier.
According to the previous analysis, the patient has the diagnosis:
DIAX
Reported on: DATEX

Your task is to determine whether the provided medical text contains a distinct instance of the diagnosis DIAX reported on a different time point rather than DATEX.

A distinct instance is defined as another occurence of the same disgnosis, but at a different time point rather than DATEX.
- a occurrence of the same diagnosis on a different date
- a clearly dated second diagnosis episode distinct from the DATEX event

Do NOT count:
- follow-up visits for the same event
- repetition of the same diagnosis without a new date
- general mentions of past history
- imaging or reports referring back to the DATEX event

Decision rules:
- If an additional instance exists, answer yes: {1}.
- If no additional instance is present, answer no: {0}.

Use the definition of the comorbidity as additional information to guide your answer:
DESCINFO

Text:
DIATEXT

Question:
Does the patient have a distinct instance of the diagnosis DIAX reported on a different time point rather than DATEX?

Allowed categories of an answer defined as ANSWERCATEGORY:
- 0
- 1

Output only the answer category in curly brackets without additional text or explanation: {ANSWERCATEGORY}.
