## Supplementary Material for "Bern Sleep-Wake Registry: temporal extraction of comorbidities from unstructured clinical text using a large language model"

### Supplementary table 1. Six comorbidity groups across 32 diagnostic categories.

| **Index** | **Diagnosis** |
| --- | --- |
| 1 | Ischemic stroke or transient ischemic attack |
| 1.1 | Transient ischaemic attack |
| 1.1.1 | Amaurosis fugax |
| 1.2 | Cerebral ischaemic stroke |
| 2.1.1 | Cerebral ischaemic stroke due to extracranial large artery atherosclerosis |
| 2.1.2 | Cerebral ischaemic stroke due to intracranial large artery atherosclerosis |
| 2.1.3 | Cerebral ischaemic stroke due to embolic occlusion |
| 2.1.4 | Cerebral ischaemic stroke due to small artery occlusion |
| 2.1.5 | Cerebral ischaemic stroke due to other known cause |
| 2 | Diabetes (any type) |
| 2.1 | Type 1 diabetes mellitus |
| 2.2 | Type 2 diabetes mellitus |
| 3 | Dyslipidemia |
| 3.1 | Hyperlipoproteinaemia |
| 3.2 | Hypercholesterolaemia |
| 3.3 | Hypertriglyceridaemia |
| 3.3.1 | Hypolipoproteinaemia |
| 4 | Hypertension |
| 4.1 | Essential hypertension |
| 4.2 | Hypertensive heart disease |
| 4.3 | Hypertensive renal disease |
| 4.4 | Pulmonary heart disease or diseases of pulmonary circulation |
| 4.4.1 | Pulmonary thromboembolism |
| 4.4.2 | Pulmonary hypertension |
| 5 | Atrial fibrillation (any type) |
| 5.1 | Paroxysmal atrial fibrillation |
| 5.2 | Persistent atrial fibrillation |
| 5.3 | Permanent atrial fibrillation |
| 6 | Sleep-disordered breathing |
| 6.1 | Central sleep apnoeas |
| 6.2 | Obstructive sleep apnoea |
| 6.3 | Sleep-related hypoventilation or hypoxemia disorders |

### Supplementary material 1. Rationale for the selection of the large language model.

Comprehensive benchmarking against six contemporary open-source models across ten diverse tasks demonstrated that gpt-oss-20b achieves mid-tier overall performance with strength in structured tasks, while consistently outperforming its larger 120B variant on several benchmarks including HumanEval and MMLU despite requiring substantially less memory and energy [10]. In clinical decision support tasks, gpt-oss-20b was shown to achieve performance comparable to or exceeding DeepSeek-R1 and o4-mini despite being substantially smaller, with fine-tuning enabling it to approach GPT-5-level diagnostic accuracy [11]. Similarly, in transplant infectious disease reasoning, the 20B variant achieved the highest correctness (81-83%) with the lowest error rate (10%), outperforming larger models and demonstrating that scale alone does not ensure clinical reliability [12]. From a deployment perspective, gpt-oss-20b delivers approximately 31.8% higher decode throughput and 25.8% lower energy per 1,000 generated tokens than Qwen3-32B, while using approximately 31.7% less peak VRAM [13].

### Supplementary material 2. Extraction prompt (in .txt).

### Supplementary material 3. Detailed description of the statistical analysis.

Statistical analyses were performed using MATLAB 2024b. LLM output was compared to the ground truth at two levels: (1) the patient level, based on information consolidated across the 2343 entries from 248 patients, and (2) the entry level, based on the individual 2343 entries. These analyses served distinct purposes. The patient-level analysis was designed to reflect the actual clinical task for information extraction (i.e., identifying the emergence of a diagnosis over the course of a patient's longitudinal record). Since a single patient could contribute multiple entries, this approach also minimized potential bias arising from repetitive entries containing semantically similar information (e.g., repeated documentation of the same diagnostic information at different time points). In contrast, the entry-level analysis enabled a more granular assessment of the LLM’s performance, including a detailed examination of the model's reasoning.

At the patient level, information from all entries belonging to the same patient was consolidated. A diagnosis was considered present if it was reported in at least one entry of the patient. Similarly, the presence of a reported date was considered positive if a date was reported in at least one entry. The questions, whether the diagnosis was present and whether the date of diagnosis was reported, were evaluated as boolean. In the statistical analysis, the question, whether the date was reported, was assessed only if the diagnosis was identified as true-positive. Agreement between the LLM output and the ground truth for these boolean outcomes was assessed using accuracy, sensitivity (recall), specificity, precision, F1 score, and Cohen’s κ. Furthermore, when a date was correctly identified as reported by both the LLM and the ground truth, date agreement was assessed. If multiple dates were available and differed in precision, the most precise date was used (e.g., 2001-06-17 instead of 2001-01-01). The analysis was done at two levels. First, agreement for the earliest reported date was evaluated by calculating the proportion of dates matching the ground truth at the day (YYYY-MM-DD), month (YYYY-MM), and year (YYYY) levels, as well as the mean and standard deviation of the difference in days between the LLM-reported and ground-truth earliest dates. The primary outcome measure was exact agreement of the earliest date at the patient level (i.e., an exact match at the day level). Second, date agreement was assessed independently of whether the number of reported dates was identical. Unique dates were compared as sets, such that a date was considered correctly identified if it was present in both the ground-truth and LLM sets. For each record, *exact-date recall* was calculated as the number of unique ground-truth dates correctly identified by the LLM divided by the total number of unique ground-truth dates in that record. The *extra-date rate* was calculated as the number of unique dates reported by the LLM that were not present in the ground truth divided by the total number of unique dates reported by the LLM for that record. *Complete exact-date agreement* was defined as equality of the ground-truth and LLM sets of unique dates, indicating that all ground-truth dates were identified and no additional dates were reported by the LLM.

At the entry level, diagnostic presence (i.e., boolean) was assessed using the approach as in the patient-level analysis. For the date analysis (i.e., down-top approach), only the most specific, leaf-level diagnostic categories were included, to avoid double-counting resulting from hierarchical aggregation of parent and child categories. Thus, when a specific end-level diagnosis was present, it was counted as one diagnostic entry, while its parent or children categories were not additionally counted. This leaf-level approach was applied to quantify the number of reported dates and their agreement. First, we reported the number of reported dates per record, including the total and mean number of dates, as well as the percentage of records with an exact match in the number of reported dates. Second, date agreement was then assessed independently of whether the number of reported dates was identical as described above.

### Supplementary material 4. Full extraction results.

### Supplementary material 5. Reasoning for “Essential hypertension” (translated into English from german original).

| Example | Reasoning |
| --- | --- |
| 1 | We need to determine if the patient has Essential hypertension. The text says: "Facteurs de risque cardiovasculaire : diabète de type 2, obésité de grade II, hypertension artérielle". That indicates hypertension arterial (blood pressure). It doesn't specify essential or secondary. But it's a risk factor; likely essential hypertension. There's no mention of secondary cause. So we can consider it as diagnosis of hypertension. The question: "Does the patient have diagnosis 'Essential hypertension'?" According to rules, if diagnosis is stated, confirmed, suspected, provisional, or listed as differential, answer {1}. Here it's a risk factor; but not explicitly essential. However, we can infer that hypertension arterial likely refers to essential hypertension unless otherwise specified. The text doesn't mention secondary cause. So we should answer {1}. Check for any rule: If diagnosis is not present or ruled-out, answer 0. It's present as "hypertension artérielle". That qualifies. There's no explicit mention of secondary cause. So yes. Thus output {1}. |
| 2 | We need to determine if the patient has Essential hypertension. The text says "arterial hypertension" which is arterial hypertension. That likely refers to essential hypertension unless secondary cause known. There's no mention of secondary cause. So we can consider it as essential hypertension. According to rules: If diagnosis stated, confirmed, suspected, provisional, or differential, answer {1}. Here it's listed under Cardiovascular risk factors: Diabetes mellitus type II, obesity grade II, arterial hypertension, obstructive sleep apnoea. So yes, essential hypertension is present. There's no mention of "secondary" or "essential". But we can infer that arterial hypertension likely refers to essential hypertension. The rule says use only information stated in text. It doesn't explicitly say "Essential hypertension", but it says "arterial hypertension". That is a diagnosis of hypertension, presumably essential unless otherwise specified. There's no mention of secondary cause. So answer {1}. |
| 3 | We need to determine if the patient has diagnosis "Essential hypertension". The text says: "Metabolic syndrome - Obesity - Arterial hypertension - Dyslipidaemia" This is German. It lists metabolic syndrome, obesity, arterial hypertension, dyslipidemia. Arterial hypertension = arterial hypertension. That likely refers to essential hypertension unless specified secondary cause. The text does not mention secondary causes or other types. So we can infer that the patient has hypertension. But question: "Does the patient have diagnosis 'Essential hypertension'?" According to rules, if diagnosis is stated, confirmed, suspected, provisional, or listed as differential, answer {1}. Here it's just a list of conditions; arterial hypertension is present. There's no mention of secondary cause. So we can consider it essential hypertension. Thus answer {1}. Check for any rule that says if not explicitly "essential" but just "arterial hypertension", we treat as essential? The definition: Essential (primary) hypertension, accounting for 95% of all cases of hypertension, is defined as high blood pressure for which a secondary cause cannot be found. Since no secondary cause mentioned, it's likely essential. Thus answer {1}. |

### Supplementary material 6. Reasoning for “Intracranial large artery atherosclerosis” (translated into English from german original).

| Example | Reasoning |
| --- | --- |
| 1 | We need to determine if patient has diagnosis "Cerebral ischaemic stroke due to intracranial large artery atherosclerosis". The text says: "Status post ischaemic brainstem infarction in the right paramedian medulla oblongata on DD.MM.YYYY - Initial symptoms: left sensorimotor hemisyndrome (NIHSS score X) - Treatment: conservative, neurological complex treatment - Aetiology: most likely microangiopathic (DD atheromatous arterial disease) - VRF: Arterial hypertension, dyslipidaemia, nicotine use, prediabetes, mild SAS (currently no indication for treatment), 3-vessel CAD - Cerebrovascular secondary diagnoses: Advanced microangiopathic leukoencephalopathy (Fazekas X)" Interpretation: The stroke is ischemic in the medulla. Etiology: "most likely microangiopathic (DD atheromatous arterial disease)". That means likely microangiopathic, differential diagnosis of atherosclerotic arterial disease. So not confirmed as large artery atherosclerosis; it's suspected but not confirmed. The question: does patient have diagnosis "Cerebral ischaemic stroke due to intracranial large artery atherosclerosis"? There's no mention of large artery atherosclerosis or specific imaging evidence. Only differential. So answer should be {0}. Also note that the text says "DD atheromatous arterial disease" meaning differential diagnosis of atherosclerotic arterial disease, but not confirmed. The rule: If diagnosis is stated, confirmed, suspected, provisional, or listed as differential, answer {1}. Wait reading decision rules: "If the diagnosis is stated, confirmed, suspected, provisional, or listed as a differential diagnosis, answer {1}." That seems to say if it's even suspected/differential, we answer 1. But that conflicts with earlier instruction: "Do not infer unsupported conditions unless there is supporting evidence directly linked to ...". The decision rules override? Let's parse carefully. The instructions at top: "Determine whether diagnosis 'Cerebral ischaemic stroke due to intracranial large artery atherosclerosis' is mentioned for the patient." Then bullet points about including confirmed, suspected, provisional, differential diagnoses if they refer to the patient. So yes, if it's suspected/differential, we should answer 1. But earlier "Do not infer unsupported conditions unless there is supporting evidence directly linked". However, suspecting atherosclerotic disease is indeed a differential; that counts as mention. The text says "DD atheromatous arterial disease" meaning differential diagnosis of atherosclerotic arterial disease. That indicates suspicion but not confirmed. So we should answer 1. But the question: "Does the patient have diagnosis 'Cerebral ischaemic stroke due to intracranial large artery atherosclerosis'?" The text says it's a differential, not confirmed. But the rule says if suspected/differential, answer {1}. So yes. Thus output {1}. Check for any contradictory statements: "most likely microangiopathic (DD atheromatous arterial disease)" means likely microangiopathic; differential is atherosclerotic arterial disease. That indicates suspicion but not confirmed. According to decision rules, that qualifies as suspected/differential => answer 1. Thus final answer {1}. |
| 2 | We need to determine if patient has diagnosis "Cerebral ischaemic stroke due to intracranial large artery atherosclerosis". The text says: "Status post ischaemic cerebral infarction in the left PICA territory on DD.MM.YYYY due to symptomatic left vertebral artery stenosis in the V4 segment - Initial symptoms: rotatory vertigo, nausea, cold sweat and oscillopsia to the left (NIHSS X) - Complications: none - Aetiology: DD arterio-arterial embolic, cardioembolic due to VHF (atrial fibrillation) - Risk factors: dyslipidaemia, sleep-related breathing disorder, atrial fibrillation - Cerebrovascular secondary diagnoses: none" Interpretation: The patient had an ischemic stroke in the PICA territory on left side. Etiology: differential diagnosis arterioarterial-embolic, cardioembolic with VHF (valvular heart disease?). Risk factors include dyslipidemia, sleep-associated breathing disorder, atrial fibrillation. No cerebrovascular comorbidities. We need to see if the stroke is due to intracranial large artery atherosclerosis. The text says "symptomatic left vertebral artery stenosis in the V4 segment". That indicates stenosis of vertebral artery (intracranial? The vertebral artery in V4 segment is intracranial portion). So there is an intracranial large artery stenosis. But the etiology is listed as differential: arterioarteriell-embolisch, cardioembolic. It doesn't explicitly state that the stroke was due to intracranial large artery atherosclerosis. The presence of vertebral stenosis suggests possible source but not confirmed. We need to see if any evidence strongly indicates diagnosis. The text says "symptomatic left vertebral artery stenosis in the V4 segment". That is an intracranial large artery atherosclerotic disease. But the stroke etiology is listed as differential: arterioarteriell-embolisch, cardioembolic. It doesn't say confirmed that it was due to vertebral stenosis. The question: "Does the patient have diagnosis 'Cerebral ischaemic stroke due to intracranial large artery atherosclerosis'?" According to decision rules: If diagnosis is stated, confirmed, suspected, provisional, or listed as differential? Wait rule says: If the diagnosis is stated, confirmed, suspected, provisional, or listed as a differential diagnosis, answer {1}. But careful: The diagnosis we are checking is "Cerebral ischaemic stroke due to intracranial large artery atherosclerosis". Is that listed as differential? The text lists etiologies: arterio-arterial embolic, cardioembolic due to VHF. It does not mention intracranial large artery atherosclerosis explicitly. So it's not listed. But the presence of vertebral stenosis might be considered evidence for intracranial large artery atherosclerosis as cause? The rule says include indirect evidence strongly indicating diagnosis: procedure performed because of that, medication clearly prescribed for that, imaging findings indicative of that, lab results indicative. Vertebral stenosis is an imaging finding indicative of intracranial large artery atherosclerosis. But does it indicate the stroke was due to that? It indicates presence of disease but not necessarily cause. The rule: "Also count indirect evidence strongly indicating diagnosis ... imaging findings indicative of 'Cerebral ischaemic stroke due to intracranial large artery atherosclerosis' (e.g., tumor on MRI from 2001 indicating tumor presence in 2001)". That example seems odd. But they say imaging findings indicative of the disease. Vertebral stenosis indicates intracranial large artery atherosclerosis. So that is indirect evidence. But does that count as diagnosis? The rule says: "If the diagnosis is stated, confirmed, suspected, provisional, or listed as a differential diagnosis, answer {1}." It doesn't say we should consider indirect evidence for diagnosing. But earlier they said include indirect evidence strongly indicating diagnosis. That suggests if there is strong indirect evidence, we can answer yes. But we need to interpret: The patient had an ischemic stroke in PICA territory; vertebral stenosis could be source. So likely the stroke was due to intracranial large artery atherosclerosis. But the text doesn't explicitly state that. However, presence of symptomatic vertebral stenosis is strong evidence for that etiology. Given guidelines: "Use only information stated in the text." The text states vertebral stenosis. That indicates intracranial large artery atherosclerosis. So we can infer that the stroke might be due to it. But they said do not infer unsupported conditions unless there is supporting evidence directly linked to diagnosis. Vertebral stenosis is directly linked. Thus answer {1}. |
| 3 | We need to determine if patient has diagnosis "Cerebral ischaemic stroke due to intracranial large artery atherosclerosis". The text: "Ischaemic cerebral infarction in the left PICA territory on DD.MM.20016 due to symptomatic left vertebral artery stenosis in the V4 segment" Interpretation: Ischemic brain infarct in left PICA territory on DD.MM.2001? Actually date maybe 2001 or 2006? It says 20016 but likely typo. The stroke is due to symptomatic vertebral stenosis at V4 segment. That is an intracranial large artery atherosclerosis (vertebral artery). So yes, the diagnosis of cerebral ischemic stroke due to intracranial large artery atherosclerosis is present. We need to answer {1}. |
| 4 | We need to determine if patient has diagnosis "Cerebral ischaemic stroke due to intracranial large artery atherosclerosis". The text says: "Status post acute ischaemic infarction in the left mesencephalon on DD.MM.YYYY - Symptoms on admission: dizziness, INO left eye (NIHSS 1) ... Aetiology: arterio-arterial embolic due to moderate-to-high-grade left vertebral artery stenosis in the V4 segment DD: microangiopathic DD: cardioembolic" So the etiology is "arterio-arterial embolic" due to moderate-high grade left vertebral artery stenosis in V4 segment. That suggests intracranial large artery atherosclerosis? The vertebral artery stenosis is an intracranial large artery (V4). So yes, it's due to intracranial large artery atherosclerosis. But the text says "arterio-arterial embolic" meaning arterial-to-arterial embolism. That could be from atherosclerotic plaque in the vertebral artery causing embolic stroke. The differential includes microangiopathic and cardioembolic. So the primary etiology is arterio-arterial embolic due to vertebral stenosis. Thus we can say diagnosis "Cerebral ischemic stroke due to intracranial large artery atherosclerosis" is present. There's no mention of ruling it out. So answer {1}. Check if any nuance: The phrase "intracranial large artery atherosclerosis" refers to a subtype of ischemic stroke classification (TOAST). The text indicates the cause is vertebral artery stenosis, which is an intracranial large artery. So yes. Thus output {1}. |

### Supplementary material 7. Reasoning for “Pulmonary thromboembolism” (translated into English from german original).

| Example | Reasoning |
| --- | --- |
| 1 | We need to determine if the patient has pulmonary thromboembolism. The text says "Status post segmental pulmonary embolism in the left upper lobe YYYY". That is a prior segmental pulmonary embolism (pulmonary embolism). So yes, diagnosis of pulmonary thromboembolism is present. It's a past event. According to rules: If the diagnosis is stated, confirmed, suspected, provisional, or listed as differential, answer {1}. So output {1}. |
| 2 | We need to determine if the patient has Pulmonary thromboembolism. The text says "Bilateral pulmonary embolism" which is bilateral pulmonary emboli (pulmonary embolism). That is pulmonary embolism. So yes, diagnosis present. Also mention of leg vein thrombosis (deep vein thrombosis) and treatment Xarelto/Eliquis. So answer {1}. |

### Supplementary material 8. Reasoning for “Hyperlipoproteinemia” (translated into English from german original).

| Example | Reasoning |
| --- | --- |
| 1 | We need to determine if the patient has Hyperlipoproteinaemia. The text is German: "Dyslipidaemia - Start Crestor due to markedly elevated Tg and LDL - no therapy of Tg". Dyslipidemia, start of Crestor due to markedly elevated triglycerides and LDL. No therapy for TG. So the patient has dyslipidemia with high TG and LDL. Hyperlipoproteinaemia is a disorder of lipoprotein metabolism resulting in high levels of lipoproteins. That includes hypercholesterolemia, hypertriglyceridemia. The text indicates elevated TG and LDL. This suggests hyperlipoproteinemia? But the question: "Does the patient have diagnosis 'Hyperlipoproteinaemia'?" According to rules: If diagnosis is stated, confirmed, suspected, provisional, or listed as differential, answer 1. Here it's not explicitly named Hyperlipoproteinaemia but dyslipidemia with elevated TG and LDL. That indicates hyperlipoproteinemia? The definition: disorders of lipoprotein metabolism that result in high levels of lipoproteins. Elevated TG and LDL qualifies. So we can consider it as evidence for hyperlipoproteinaemia. There's no mention of ruling out. So answer {1}. |
|  | We need to determine if the provided medical text mentions that the patient has a diagnosis "Hyperlipoproteinaemia". The text is: "Statin intolerancexa0 xa0 - Symmetric typical myopathy of the proximal thigh and upper arm muscles xa0 - On Repatha" This appears to be German: "Statin intolerance - symmetric typical myopathy of the proximal thigh and upper arm muscles - under Repatha". So patient has statin intolerance, myopathy. No mention of hyperlipoproteinaemia. There's no mention of medication for hyperlipoproteinaemia? Repatha is a PCSK9 inhibitor used to lower LDL cholesterol; it's prescribed for hyperlipidemia or high LDL. But the text says "On Repatha" meaning "under Repatha". That indicates they are on Repatha, which is used for hyperlipoproteinemia (high lipids). However, the question: "Does the patient have diagnosis 'Hyperlipoproteinaemia'?" The presence of medication could be indirect evidence strongly indicating diagnosis. According to instructions: include indirect evidence strongly indicating diagnosis of "Hyperlipoproteinaemia", including medication clearly prescribed for "Hyperlipoproteinaemia" (e.g., lisinopril for arterial hypertension). Wait, that example is wrong: lisinopril is for hypertension not hyperlipidemia. But they say medication clearly prescribed for Hyperlipoproteinaemia. Repatha is indeed used to treat high LDL cholesterol, i.e., hyperlipoproteinemia. So presence of Repatha indicates the patient has hyperlipoproteinemia. But we must be careful: The instructions mention "medication clearly prescribed for 'Hyperlipoproteinaemia' (e.g., lisinopril for arterial hypertension)". That example is wrong but they intend to say medication used for that condition. So yes, Repatha indicates hyperlipoproteinemia. Thus the answer should be {1}. Check if any other evidence: statin intolerance suggests high cholesterol requiring alternative therapy like PCSK9 inhibitors. So yes. Therefore output {1}. |
|  | We need to determine if the patient has diagnosis Hyperlipoproteinaemia. The text includes "Atorvastatin 40mg" medication, which is a statin used for hyperlipidemia/hypercholesterolemia. But does that count as evidence of diagnosis? According to instructions: medication clearly prescribed for Hyperlipoproteinaemia counts as indirect evidence strongly indicating diagnosis. So yes, Atorvastatin indicates treatment for high lipids. Also "Arterial hypertension" is listed but not relevant. Also the patient has "Substituted hypothyroidism" etc. No mention of hyperlipidemia explicitly. But medication suggests it. The instructions: include indirect evidence strongly indicating diagnosis, including medication clearly prescribed for Hyperlipoproteinaemia (e.g., lisinopril for arterial hypertension). Wait that example is wrong? They gave lisinopril for arterial hypertension as an example of medication for hypertension, not hyperlipidemia. But they say "medication clearly prescribed for 'Hyperlipoproteinaemia' (e.g., lisinopril for arterial hypertension)". That seems inconsistent: lisinopril is antihypertensive, not hyperlipidemia. Maybe a mistake. But we can interpret that medication used to treat hyperlipidemia counts. Atorvastatin is definitely used for hyperlipidemia. So answer {1}. Check if any rule says if diagnosis not present or ruled-out: no mention of "no Hyperlipoproteinaemia". So we go with 1. Thus output {1}. |

### Supplementary material 9. Reasoning for “Sleep-related hypoventilation or hypoxemia disorders” (false-negatives; translated into English from german original).

| Example | Reasoning |
| --- | --- |
| 1 | We need to determine if patient has diagnosis "Sleep-related hypoventilation or hypoxemia disorders". The text: Severe obstructive sleep apnoea, ED MM/YYYY - MM/YYYY PSG: AHI X/h, hypopnoeas X/h, oAI X/h, cAI X/h, mixed apnoea X/h, ODI X/h, mean tcCO2 46mmHg, maximum tcCO2 most likely 55mmHg, mean pCO2 93% We have data: AHI high, ODI X/h (oxygen desaturation index). Also mention of "mean tcCO2 46 mmHg" and "maximum tcCO2 most likely 55 mmHg", "mean pCO2 93%". Wait pCO2 93%? That seems odd; maybe it's 93 mmHg? But anyway, they have elevated CO2 during sleep. The question: Does the patient have diagnosis "Sleep-related hypoventilation or hypoxemia disorders"? The definition requires objective evidence of hypercapnia (PaCO2) or sustained oxygen desaturation. Here we have ODI X/h indicates frequent desaturations; also mention of elevated CO2 levels during sleep. That suggests hypoventilation. But the rule: "Answer {1} ONLY if the text explicitly mentions hypoventilation, hypercapnia, or hypoxemia during sleep." The text includes "mean tcCO2 46mmHg" and "maximum tcCO2 most likely 55mmHg", "mean pCO2 93%". That indicates elevated CO2 (hypercapnia). Also ODI high suggests hypoxemia. Thus we can answer {1}. Check if any mention of "Sleep-related hypoventilation or hypoxemia disorders" explicitly? No, but the evidence qualifies. So answer {1}. |
| 2 | We need to determine if patient has diagnosis "Sleep-related hypoventilation or hypoxemia disorders". The text includes polysomnography data. It shows ODI (oxygen desaturation index) high, SpO2 average 90.1% and 88%. Also SpO2 < 90% for 185 min in one study. But does it mention sustained decline to ≤88% for ≥5 minutes? They have SpO2 < 90% 185 min. That indicates hypoxemia during sleep. The definition: Sleep-related hypoxemia diagnosed when overnight monitoring reveals sustained (>5 min) decline in oxygen saturation to ≤88% (or ≤90% for children). Here we have SpO2 < 90% for 185 minutes, but not necessarily sustained >5 min at ≤88%. But average SpO2 88%, and SpO2 < 90% 185 min. That suggests hypoxemia. Also ODI high indicates desaturations. Also mention of "mtcCO2 38 mmHg, max 44 mmHg" - normal CO2? Not hypercapnia. So no hypoventilation evidence. But hypoxemia is present. The question: does the patient have diagnosis "Sleep-related hypoventilation or hypoxemia disorders"? According to rules: answer {1} only if text explicitly mentions hypoventilation, hypercapnia, or hypoxemia during sleep. Here we have hypoxemia evidence: SpO2 < 90% for 185 min, average SpO2 88%. That qualifies as sustained decline? It's not clear if it's sustained >5 minutes at ≤88%, but they had SpO2 < 90% for 185 min. The definition says sustained (>5 min) decline to ≤88% in adults. They have average 88% and SpO2 < 90% for 185 min, but we don't know if it was below 88%. But the presence of ODI 87.2/h indicates many desaturations. Also "SpO2 < 90% 185 min" suggests hypoxemia. Thus likely yes. The question: "Does the patient have diagnosis 'Sleep-related hypoventilation or hypoxemia disorders'?" According to rules, answer {1} if text explicitly mentions hypoxemia during sleep. It does mention SpO2 < 90% for 185 min and average SpO2 88%. That indicates hypoxemia. Thus output {1}. |
| 3 | We need to determine if patient has diagnosis "Sleep-related hypoventilation or hypoxemia disorders". The text says: "Moderate obstructive sleep apnoea and nocturnal alveolar hypoventilation - DD in the context of chronic sensorimotor axonal demyelinating polyneuropathy and reduced vocal cord mobility - PSG DD.MM.YYYY: AHI X/h OSA, AASM hypoventilation criteria just met, mean oxygen saturation X%, ODI X/h" So it states "nocturnal alveolar hypoventilation" (nighttime alveolar hypoventilation). Also says "AASM hypoventilation criteria just met". That indicates hypoventilation during sleep. The question: does the patient have diagnosis "Sleep-related hypoventilation or hypoxemia disorders"? According to rules, answer {1} only if text explicitly mentions hypoventilation, hypercapnia, or hypoxemia during sleep. Here it says alveolar hypoventilation and AASM criteria fulfilled. So yes. Thus output {1}. |
| 4 | We need to determine if the patient has diagnosis "Sleep-related hypoventilation or hypoxemia disorders". The text is German. It says: "Moderate obstructive sleep apnoea and nocturnal alveolar hypoventilation - DD in the context of chronic sensorimotor axonal demyelinating polyneuropathy and reduced vocal cord mobility - Therapy: BiPAP training by pulmonology planned" Translation: "Moderate obstructive sleep apnea and nocturnal alveolar hypoventilation – differential diagnosis in the context of chronic sensomotoric axonal demyelinating polyneuropathy and vocal fold paresis – therapy: BiPAP training by pulmonology pla ed." So it explicitly mentions "nocturnal alveolar hypoventilation" which is nocturnal alveolar hypoventilation. That is a form of sleep-related hypoventilation. The question: Does the patient have diagnosis "Sleep-related hypoventilation or hypoxemia disorders"? According to rules, answer {1} if text explicitly mentions hypoventilation, hypercapnia, or hypoxemia during sleep. Yes, it says nocturnal alveolar hypoventilation. So it's present. Thus output {1}. |
| 5 | We need to determine if patient has diagnosis "Sleep-related hypoventilation or hypoxemia disorders". The text says: "nocturnal hypercapnia - most likely artefactual - Polysomnography DD.MM.YYYY: pCO2 48-56 mmHg" So nocturnal hypercapnia, artifactually? It says "most likely artefactual" maybe artifactually? But the key is that polysomnography shows pCO2 48-56 mmHg. That indicates elevated CO2 during sleep. The definition of Sleep-related hypoventilation: abnormally elevated PaCO2 during sleep. So yes, this is evidence. We need to answer {1} because diagnosis is present (confirmed). There's no mention of "hypoxemia" but hypercapnia qualifies as hypoventilation. So answer {1}. |
| 6 | We need to determine if patient has diagnosis "Sleep-related hypoventilation or hypoxemia disorders". The text says: "Severe mixed sleep apnoea:" (severe mixed sleep apnea). PSG MM/YYYY: AHI X/h, ODI X/h, predominantly hypopneas, about X% with central character, central apneas X/h, mixed apneas X/h. Arousal index X/h, of which respiratory arousals X/h (spontaneousX/h). - Nighttime hypoxemia: SpO2 <90% for 5 minutes, mean SpO2 93%. Clinical: fatigue, concentration disturbances, forgetfulness. Risk factor: BMI XX. We need to see if the patient has "Sleep-related hypoventilation or hypoxemia disorders". The definition says: primary feature is insufficient sleep related ventilation leading to elevated PaCO2 during sleep. Sleep-related hypoxemia diagnosed when overnight monitoring reveals sustained (>5 min) decline in oxygen saturation to ≤88% (or ≤90% for adults?) Actually the text says "≤ 88%" but also note that for adults it's ≤88%. But the text says SpO2 <90% for 5 minutes. That is a drop below 90% for 5 minutes, but not necessarily below 88%. The definition: sustained (>5 min) decline in oxygen saturation to ≤ 88% in adults (or ≤90% in children). So for adults, threshold is 88%. But the text says SpO2 <90% for 5 minutes. That might be considered hypoxemia if it goes below 90%? The definition says "≤ 88%" but also note that some guidelines use ≤90% for children. However, the question: does the patient have diagnosis "Sleep-related hypoventilation or hypoxemia disorders"? We need explicit mention of hypoventilation, hypercapnia, or hypoxemia during sleep. The text says nighttime hypoxemia: SpO2 <90% 5 minutes, mean SpO2 93%. That indicates a drop below 90% for at least 5 minutes. But the definition threshold is ≤88% for adults. However, the question might accept that "SpO2 <90%" qualifies as hypoxemia? The instructions: "Answer {1} ONLY if the text explicitly mentions hypoventilation, hypercapnia, or hypoxemia during sleep." It says nighttime hypoxemia: SpO2 <90% 5 minutes. That is a mention of hypoxemia (SpO2 drop). So yes, it qualifies as hypoxemia. Even though threshold might be 88%, the text explicitly states "SpO2 <90%" for 5 min. The question says "explicitly mentions hypoventilation, hypercapnia, or hypoxemia during sleep." It does mention nighttime hypoxemia. So answer {1}. We should also consider that mixed apnea includes central and obstructive components but not necessarily hypoventilation. But the presence of nighttime hypoxemia qualifies as "Sleep-related hypoxemia disorders". So we answer {1}. |
| 7 | We need to determine if patient has diagnosis "Sleep-related hypoventilation or hypoxemia disorders". The text includes polysomnography results with oxygen desaturations. It says Sp02 <90% for 5.5 min in MM/YYYY, and Sp02 <90% for 16.5 min in MM/YYYY. That meets criteria for sleep-related hypoxemia: sustained decline to ≤88% for ≥5 minutes. They have desaturations down to 74-75%. So yes, they meet objective evidence of sleep-related hypoxemia. Also mention "No evidence of hypoventilation (mean tpCO2 40.9 mmHg)" meaning no evidence of hypoventilation. But the question is about diagnosis "Sleep-related hypoventilation or hypoxemia disorders". That includes either hypoventilation OR hypoxemia. Since they have hypoxemia, that qualifies. Thus answer {1}. Check rule: If diagnosis explicitly mentioned? Not necessarily; but we can infer from evidence. The rule says: Answer {1} ONLY if text explicitly mentions hypoventilation, hypercapnia, or hypoxemia during sleep. Here it does mention Sp02 <90% for 5.5 min and 16.5 min. That indicates hypoxemia. So yes. Thus output {1}. |
| 7 | We need to determine if patient has diagnosis "Sleep-related hypoventilation or hypoxemia disorders". The text: "Suspected obstructive sleep apnoea syndrome - Epworth Sleepiness Scale XX/24 points - Pulse oximetry DD.MM.YYYY: AHI X/h, ODI X/h, SpO2 min. 79%" So they have obstructive sleep apnea syndrome (OSA). They mention SpO2 minimum 79%. That indicates hypoxemia during sleep? The definition says Sleep-related hypoxemia is diagnosed when overnight monitoring reveals sustained (>5 minutes) decline in oxygen saturation to ≤88% for ≥5 minutes. Here we only see a single measurement of min SpO2 79%, but not duration. But the presence of ODI 11/h indicates desaturation events >3%? ODI stands for Oxygen Desaturation Index: number of times per hour that SpO2 drops by at least 3%. So there are 11 events per hour, indicating repeated hypoxemia episodes. But the question: "Does the patient have diagnosis 'Sleep-related hypoventilation or hypoxemia disorders'?" According to rules: answer {1} only if text explicitly mentions hypoventilation, hypercapnia, or hypoxemia during sleep. Here we have SpO2 min 79% and ODI X/h. That indicates hypoxemia episodes. So yes, evidence of hypoxemia. But the rule says: "Answer {1} ONLY if the text explicitly mentions hypoventilation, hypercapnia, or hypoxemia during sleep." The text includes SpO2 min 79%. That's a measure of oxygen saturation during sleep. It indicates hypoxemia. So we can answer {1}. There's no mention of hypercapnia or CO2. Thus output: {1}. |
| 9 | We need to determine if patient has diagnosis "Sleep-related hypoventilation or hypoxemia disorders". The text includes polysomnography data: AHI high, mean tcCO2 46 mmHg, max 50 mmHg. That indicates hypercapnia during sleep (mean transcutaneous CO2). Sleep-related hypoventilation is defined by elevated PaCO2 during sleep. So yes, evidence of hypercapnia. Also mention "SARS CoV2 screening" irrelevant. Thus answer {1}. |
| 10 | We need to determine if patient has diagnosis "Sleep-related hypoventilation or hypoxemia disorders". The text is German. Let's translate key parts: "Central breathing disorder in the context of Joubert syndrome - Daytime hyperventilation episodes - At night central sleep apnoea syndrome, severe, with: - Polysomnography MM/ YYYY: AHI X/h, cAI X/h, hypopnoeas with central character X/h, phases of hyperventilation. TcpCO2 between 40 and 52.9mmHg (time above 50 mmHg 148 minutes). Obstructive apnoea index X/h. Oxygen desaturation index X/h. Mean oxygen saturation 97.4%, minimum 93%. - Polysomnography MM/YYYY: persistent central sleep apnoea (AHI X/h, cAI X/h) and phases of hyperventilation (respiratory rate X/h vs. X/h)." So we have central sleep apnea with hyperventilation phases. TcpCO2 between 40 and 52.9 mmHg; time over 50 mmHg 148 minutes. That indicates elevated CO2 during sleep? The definition: Sleep-related hypoventilation is insufficient ventilation leading to abnormally elevated PaCO2 during sleep. Here we have increased CO2 >50 mmHg for some time, which suggests hypercapnia. Also mention of hyperventilation episodes. But the question: "Does the patient have diagnosis 'Sleep-related hypoventilation or hypoxemia disorders'?" We need explicit mention of hypoventilation/hypercapnia/hypoxemia during sleep. The text says "TcpCO2 between 40 and 52.9 mmHg (time over 50 mmHg 148 minutes)". That indicates hypercapnia >50 mmHg for some time, which is a sign of hypoventilation. Also mention of hyperventilation episodes but that might be paradoxical. Also oxygen saturation: minimal 93%, average 97.4%. No sustained drop to ≤88% for ≥5 min. So no hypoxemia evidence. But we have hypercapnia >50 mmHg for 148 minutes, which is more than 5 minutes? Yes, 148 minutes >5 min. That indicates sleep-related hypoventilation. Thus the diagnosis "Sleep-related hypoventilation or hypoxemia disorders" is present (confirmed). So answer {1}. Check rule: Must explicitly mention hypoventilation/hypercapnia/hypoxemia during sleep. The text mentions elevated CO2 >50 mmHg for 148 minutes, which qualifies as hypercapnia during sleep. That indicates hypoventilation. So yes. Thus output {1}. |

### Supplementary material 10. Reasoning for “Sleep-related hypoventilation or hypoxemia disorders” (false-positives; translated into English from german original).

| Example | Reasoning |
| --- | --- |
| 1 | We need to determine if patient has diagnosis "Sleep-related hypoventilation or hypoxemia disorders". The text mentions: "moderate restrictive ventilatory disorder with respiratory insufficiency (BiPAP therapy) since age X" So the patient has restrictive ventilatory disorder with respiratory insufficiency, BiPAP therapy since age X. That indicates sleep-related hypoventilation? The definition: Sleep-related hypoventilation is insufficient ventilation during sleep leading to hypercapnia. BiPAP therapy suggests treatment for hypoventilation or OSA. But the text does not explicitly mention hypoventilation, hypercapnia, or hypoxemia during sleep. It says "restrictive ventilatory disorder with respiratory insufficiency". That could be due to neuromuscular disease causing hypoventilation. BiPAP therapy is used for hypoventilation disorders. But the rule: For diagnosis "Sleep-related hypoventilation or hypoxemia disorders", answer {1} ONLY if text explicitly mentions hypoventilation, hypercapnia, or hypoxemia during sleep. Answer {0} if only mentions apnoea, OSA, CSA, SDB without these terms. Thus we need explicit mention of hypoventilation/hypercapnia/hypoxemia during sleep. The text does not explicitly say "hypoventilation" but says "restrictive ventilatory disorder with respiratory insufficiency". That is a ventilatory disorder, but not necessarily sleep-related. However BiPAP therapy indicates treatment for hypoventilation or OSA. But the rule says we need explicit mention of hypoventilation/hypercapnia/hypoxemia during sleep. The text does not say "sleep" anywhere. So by strict rule, answer {0}. But maybe we consider that BiPAP therapy is used for sleep-related hypoventilation or OSA. But the rule says only if explicitly mentions hypoventilation etc. It doesn't mention "sleep". So I'd go with {0}. |
| 2 | We need to determine if patient has diagnosis "Sleep-related hypoventilation or hypoxemia disorders". The text says: "obstructive sleep apnoea (ventilation device not tolerated";" on nocturnal oxygen for diagnosis 2)". That indicates obstructive sleep apnea. No mention of hypoventilation, hypercapnia, hypoxemia during sleep. There's mention of nocturnal oxygen therapy. But that is not necessarily evidence of hypoxemia? The text says "on nocturnal oxygen" meaning under nocturnal oxygen at diagnosis 2. That could be due to OSA or other reasons. However the rule: answer {1} only if text explicitly mentions hypoventilation, hypercapnia, or hypoxemia during sleep. It does not mention those terms. So answer {0}. Also no mention of polysomnography with CO2 monitoring. So {0}. |

### Supplementary material 11. Reasoning for additional instances of “Essential hypertension” (translated into English from german original).

| Example | Reasoning |
| --- | --- |
| 1 | We need to parse the text. The diagnosis is "Essential hypertension" (essential hypertension). We have a date MM/YYYY, then MM/YYYY, then DD.MM.YYYY, and DD.MM.YYYY. The question: does patient have distinct instance of diagnosis reported on different time point than DD.MM.YYYY? Yes, there are later dates with hypertension mention. We need to check if the text includes a distinct occurrence of essential hypertension at a different date. The text says "Inadequate blood pressure control: hypertension grade 3" but no explicit date attached except earlier context maybe MM/YYYY or DD.MM.YYYY? Let's parse: Text snippet: "Essential hypertension and hypertensive cardiopathy - MM/YYYY renal denervation - MM/YYYY TTE: Normal syst. LV and RV function"; concentric LV hypertrophy with diastolic dysfunction;" no relevant valvular heart disease - Inadequate blood pressure control: hypertension grade X - Plasma levels of aldosterone, renin and metanephrines within normal range - DD.MM.YYYY 24h blood pressure measurement: BP values over 24h XXX/XX mmHg, systol. dipping X% Currently: - DD.MM.YYYY TTE: ..." So the diagnosis of essential hypertension appears at MM/YYYY (first mention). Then later there is "Inadequate blood pressure control: hypertension grade X" but no date; likely part of MM/YYYY? Actually the bullet list: "- MM/YYYY renal denervation - MM/YYYY TTE: Normal syst. LV and RV function"; ... "Inadequate blood pressure control: hypertension grade 3" maybe still under MM/YYYY? Hard to parse but likely the hypertension is ongoing. Then DD.MM.YYYY 24h blood pressure measurement shows high values, indicating hypertension again. So that is a distinct instance at a different date (MM YYYY). Also DD.MM.YYYY current status includes TTE etc. But no explicit mention of hypertension but likely still present. Thus answer yes: {1}. |
| 2 | Essential hypertension" reported on DD.MM.YYYY and DD.MM.YYYY. We must determine if there is a distinct instance of the same diagnosis at a different time point in the provided text. The text: "Essential hypertension and hypertensive cardiopathy - 0 MM/YYYY renal denervation - MM/YYYY TTE: Normal syst. LV and RV function"; concentric LV hypertrophy with diastolic dysfunction;" no relevant valvular heart disease - Inadequate blood pressure control: hypertension grade 3 - Plasma levels of aldosterone, renin and metanephrines within normal range - DD.MM.YYYY 24h blood pressure measurement: BP values over 24h XXX/XX mmHg, systol. dipping XX% Currently: - DD.MM.YYYY TTE: Normal systolic LV and RV function, concentric LV hypertrophy with diastolic dysfunction, no relevant valvular heart disease. - DD.MM.YYYY Cardiopulmonary exercise test: ..." We need to see if there's a distinct instance of "Essential hypertension" at a different date than the two given dates (DD.MM.YYYY and DD.MM.YYYY). The text includes "Essential hypertension and hypertensive cardiopathy - MM/YYYY renal denervation". That is MM YYYY. Also "DD.MM.YYYY 24h blood pressure measurement: BP values over 24h XXX/XX mmHg" indicates blood pressure measurement but not explicitly diagnosis of essential hypertension, though it says "Inadequate blood pressure control: hypertension grade X". That is a mention of hypertension grade X. But the question: distinct instance of the diagnosis "Essential hypertension" reported on a different time point. We have at least one date: MM/YYYY (MM YYYY). Also DD.MM.YYYY measurement, but not explicitly labeled as essential hypertension; it's just blood pressure values and mention of hypertension grade X. But the diagnosis "Essential hypertension" appears in the text with date MM/YYYY. That is a distinct instance from DD.MM.YYYY? Wait DD.MM.YYYY is MM X, YYYY. The text says MM YYYY renal denervation. That likely refers to MM YYYY but not necessarily the exact day. But it's still a different date point (MM YYYY). However we need distinct instance of diagnosis "Essential hypertension" reported on a different time point than DD.MM.YYYY and DD.MM.YYYY. We have MM/YYYY, which is MM YYYY. That could be considered the same month but not necessarily the exact day. But it's still a date point distinct from DD.MM.YYYY? It might be ambiguous: MM/YYYY could refer to any day in MM YYYY; if we consider it as MM X, YYYY, then it's the same date. But likely it's just month/year. The question says "different time point rather than DD.MM.YYYY, DD.MM.YYYY." So any other date qualifies. Thus yes, there is a distinct instance: DD.MM.YYYY measurement indicates hypertension grade X; but the diagnosis of essential hypertension might be implied. But we need explicit mention "Essential hypertension" or "Essential hypertension". The text includes "Essential hypertension and hypertensive cardiopathy - MM/YYYY renal denervation". That is a distinct instance at MM YYYY. Thus answer: yes => {1}. But we must be careful: The question says "distinct instance of the diagnosis 'Essential hypertension' reported on a different time point rather than DD.MM.YYYY, DD.MM.YYYY." So if there is any mention of essential hypertension at a date other than those two dates. We have "Essential hypertension" in MM YYYY. That qualifies. Thus answer: {1}. |
| 3 | We need to parse the text. The diagnosis "Essential hypertension" is present on DD.MM.YY (TTE of DD.MM.YY). Also mention of Renal denervation on DD.MM.YY. Then " MM YYYY: inadequate blood pressure control hypertension grade 2". That indicates a different time point, MM YYYY, with hypertension grade X. Is that considered a distinct instance? The question: "Does the patient have a distinct instance of the diagnosis 'Essential hypertension' reported on a different time point rather than DD.MM.YYYY?" We need to see if there's an occurrence of same diagnosis at a different date. The text says " MM YYYY: inadequate blood pressure control hypertension grade X". That is a mention of hypertension grade X. Is that the same diagnosis? It's still hypertension, but maybe it's not explicitly labeled as "Essential hypertension" (essential). But it might be considered a distinct instance because it's a different date and mentions hypertension. But we need to check if it's specifically "Essential hypertension". The text: "Hypertensive cardiopathyxa0 - Essential hypertension xa0 - TTE of DD.MM.YY..." So the diagnosis is "Essential hypertension" (essential hypertension). Then later: " MM YYYY: inadequate blood pressure control hypertension grade X". That might be a separate mention of hypertension but not explicitly labeled as essential. However, it's likely still essential hypertension because no secondary cause is mentioned. But the question: "Does the patient have a distinct instance of the diagnosis 'Essential hypertension' reported on a different time point rather than DD.MM.YYYY?" The text includes a MM YYYY mention of hypertension grade X. That could be considered a distinct instance, but it's not explicitly labeled as essential. But we can infer that it's still essential because no secondary cause is found. But the instructions: "Use the definition of the comorbidity as additional information to guide your answer." The definition says essential hypertension is high blood pressure for which a secondary cause cannot be found. There's no mention of secondary causes in MM YYYY, so likely it's essential. Thus we have an additional instance on MM YYYY. So answer yes: {1}. Check if any other date? Only DD.MM.YY and MM YYYY. So distinct instance exists. So output {1}. |

### Supplementary material 12. Reasoning for additional instances of “Sleep-related hypoventilation or hypoxemia disorders” (translated into English from german original).

| Example | Reasoning |
| --- | --- |
| 1 | We need to determine if there's a distinct instance of diagnosis "Sleep-related hypoventilation or hypoxemia disorders" reported on a different time point than YYYY -MM-DD. The patient has known sleep-related hypoventilation since YYYY, and there is PSG MM/YY: Sleep-related hypoventilation (Polysomnography MM YYYY). Also "nocturnal hypoventilation, known since YYYY, non-invasive ventilation since autumn YYYY ". So the diagnosis was known in YYYY. The question: does the patient have a distinct instance reported on a different time point? Yes, PSG MM/YY indicates a new evaluation of sleep-related hypoventilation at MM YYYY. That is a distinct occurrence with objective evidence. So answer yes: {1}. |
| 2 | We need to determine if there's a distinct instance of diagnosis "Sleep-related hypoventilation or hypoxemia disorders" reported on a different time point than YYYY-MM-DD and YYYY-MM-DD. The provided text includes: - PSG MM/YY: Sleep-related hypoventilatio - nocturnal hypoventilation, known since YYYY, non-invasive ventilation since autumn YYYY, meanwhile better compliance in this regard for approx. X months So we have a diagnosis of sleep-related hypoventilation reported in PSG MM/YY (November YYYY). That is a different date from YYYY-MM-DD and YYYY-MM-DD. So yes, there is an additional instance. Thus answer: {1}. |

### Supplementary material 13. Reasoning for additional instances of “Sleep-related hypoventilation or hypoxemia disorders” (translated into English from german original).

| Example | Reasoning |
| --- | --- |
| 1 | The document states “MM/YYYY” indicating the polysomnography was performed in MM YYYY, which is represented by the extracted date YYYY-MM-DD. |
| 2 | The text indicates a wake‑up stroke occurring on DD.MM.YYYY, matching the extracted date. |
| 3 | The evaluation date is indicated as September YYYY (ED MM/YYYY), so YYYY-MM-DD appropriately represents the month when no exact day is provided. |
| 4 | The text indicates that type 2 diabetes was diagnosed around YYYY, so the extracted date of YYYY-MM-DD appropriately reflects that approximate year. |
| 5 | The extracted date YYYY-MM-DD corresponds to the documented wake‑up stroke on YYYY-MM-DD, during which dyslipidemia is listed as a vascular risk factor. |
